# A combined risk model shows viability for personalized breast cancer risk assessment in the Indonesian population

**DOI:** 10.1101/2023.09.22.23295602

**Authors:** Bijak Rabbani, Sabrina Gabriel Tanu, Kevin Nathanael Ramanto, Jessica Audrienna, Fatma Aldila, Eric Aria Fernandez, Mar Gonzalez-Porta, Margareta Deidre Valeska, Jessline Haruman, Lorina Handayani Ulag, Yusuf Maulana, Kathleen Irena Junusmin, Margareta Amelia, Gabriella Gabriella, Feilicia Soetyono, Aulian Fajarrahman, Salma Syahfani Maudina Hasan, Faustina Audrey Agatha, Marco Wijaya, Stevany Tiurma Br Sormin, Levana Sani, Soegianto Ali, Astrid Irwanto, Samuel J Haryono

## Abstract

Breast cancer remains a significant concern worldwide, with a rising incidence in Indonesia. This study aims to evaluate the applicability of risk-based screening approaches in the Indonesian demographic through a case-control study involving 305 women. We developed a personalized breast cancer risk assessment workflow that integrates multiple risk factors, including clinical (Gail) and polygenic (Mavaddat) risk predictions, into a consolidated risk category. By evaluating the area under the receiver operating characteristic curve (AUC) of each single-factor risk model, we demonstrate that they retain their predictive accuracy in the Indonesian context (AUC for clinical risk: 0.67 [0.61,0.74]; AUC for genetic risk: 0.67 [0.61,0.73]). Notably, our combined risk approach enhanced the AUC to 0.70 [0.64,0.76], highlighting the advantages of a multifaceted model. Our findings demonstrate for the first time the applicability of the Mavaddat and Gail models to Indonesian populations, and show that within this demographic, combined risk models provide a superior predictive framework compared to single-factor approaches.

## Introduction

Breast cancer is a pressing global public health concern. It has emerged as the most commonly diagnosed type of cancer worldwide, with its prevalence steadily rising in recent years. For example, in 2020 alone, an estimated 2.48 million cases were reported worldwide, resulting in approximately 685,000 deaths^1^. In Indonesia, breast cancer mirrors the global trajectory, establishing itself as a leading cause of mortality among women^2^. Specifically, in 2020, GLOBOCAN documented over 68,000 new diagnoses and 22,000 resultant deaths, representing incidence and mortality rates of 15.3% and 11%, respectively^3^. Thus, addressing the burden of breast cancer is crucial for improving public health outcomes.

As with other cancers, prompt diagnosis and timely therapeutic interventions have been unequivocally associated with enhanced patient prognosis and diminished mortality rates. Specifically, survival rates for stage 1 breast cancer are estimated at 99% five years post-diagnosis, but this figure quickly diminishes to 30% for advanced stages^4^. As a result, numerous countries have established population screening programs to promote early detection of breast cancer. Mammography remains the primary method for breast cancer screening and has been proven to decrease mortality rates^5,6^. Nonetheless, mammography is not devoid of challenges. Typically, it adopts a uniform approach, recommending all women within a certain age bracket to undergo the procedure^6^. Moreover, in Asian populations, women present higher breast tissue density, which complicates mammogram interpretation and can lead to false-positive diagnoses^7^. Compounding the challenges above, Indonesia has yet to incorporate breast cancer screening into its national health agenda, and the procedure remains unsupported by the national health insurance framework. This financial constraint dissuades women from pursuing mammographic screening and compromises diagnostic rates when the disease is the most actionable^8^. For instance, a longitudinal analysis conducted over three decades at multiple academic hospitals found that a significant proportion of patients, ranging from 68-73%, only sought medical consultations during the advanced stages of the disease^2^.

Collectively, the observations highlighted above underscore the need for refining breast cancer screening strategies, emphasizing both accessibility and diagnostic precision. Recently, risk-based screening, which customizes recommendations according to individual risk profiles, has challenged the traditional one-size-fits-all screening paradigms, and is emerging as a promising approach for enhanced patient stratification^9–12^. A myriad of risk factors, encompassing clinical predispositions, familial history, and genetic markers, have been considered into these advanced screening methods. For example, the Gail model, a non-genetic risk assessment tool, offers insights into the probability of an individual developing breast cancer over a five-year span based on clinical risk factors, including age, reproductive history, and familial breast cancer incidence^13^. Similarly, pathogenic mutations in high-penetrance genes, such as BRCA1 and BRCA2, which are involved in DNA repair, have long been recognised as pivotal risk determinants for breast cancer and have been incorporated into routine clinical practice^14^. More recently, Polygenic Risk Scores (PRS) such as the Mavaddat model, which aggregate the effects of multiple low-penetrance genes, have been demonstrated to harbour predictive power comparable to their high-penetrance counterpart^15,16^. However, despite advances in risk-based screening, most research and model development has been focused on Western populations^17^. This leaves a gap in understanding the applicability of these models to Southeast Asian populations, such as Indonesia. Unique genetic, environmental, and clinical factors may affect breast cancer risk in these populations, so validating and adapting these models is essential.

In the present study, we assess the applicability of two notable breast cancer risk assessment models to the Indonesian demographic, based on a case-control study involving 305 women. We focus on the predictive accuracy of the Gail model, which evaluates clinical risk, the Mavaddat model, which assesses polygenic risk, as well as pathogenic mutations in BRCA1/2 genes, which are representative of monogenic risk. Additionally, we utilize a combined risk model to categorise patients into either elevated or average risk groups. Our objective is to address the current knowledge gap and offer a risk assessment instrument specifically designed for the Indonesian context.

## Results

### Development and validation of a personalised breast cancer risk assessment workflow

We have developed a personalized breast cancer risk assessment workflow that integrates data from multiple risk factors, including assessment of clinical risk based on the Gail model, evaluation of polygenic risk through the Mavaddat model, and monogenic risk through detection of pathogenic/likely pathogenic mutations in BRCA1/2 (**Figure 1**). The process begins with a pre-test counselling session, during which eligible participants, upon providing informed consent, submit a buccal sample and complete a risk survey. This sample is then processed in a testing laboratory where it is genotyped using microarrays. Bioinformatic analyses are next employed to calculate ancestry-adjusted PRS and to translate these into 5-year absolute risk scores, leveraging localized breast cancer incidence and mortality data. Array results are also inspected to determine the presence of pathogenic/likely pathogenic mutations in BRCA1/2 genes. Lastly, responses from the clinical risk survey are analysed to derive 5-year absolute risk scores in accordance with the Gail model. In a final step, both the genetic and clinical risk scores are combined into a risk category, with participants receiving individualized risk reports during a post-test consultation.

**Figure 1:**
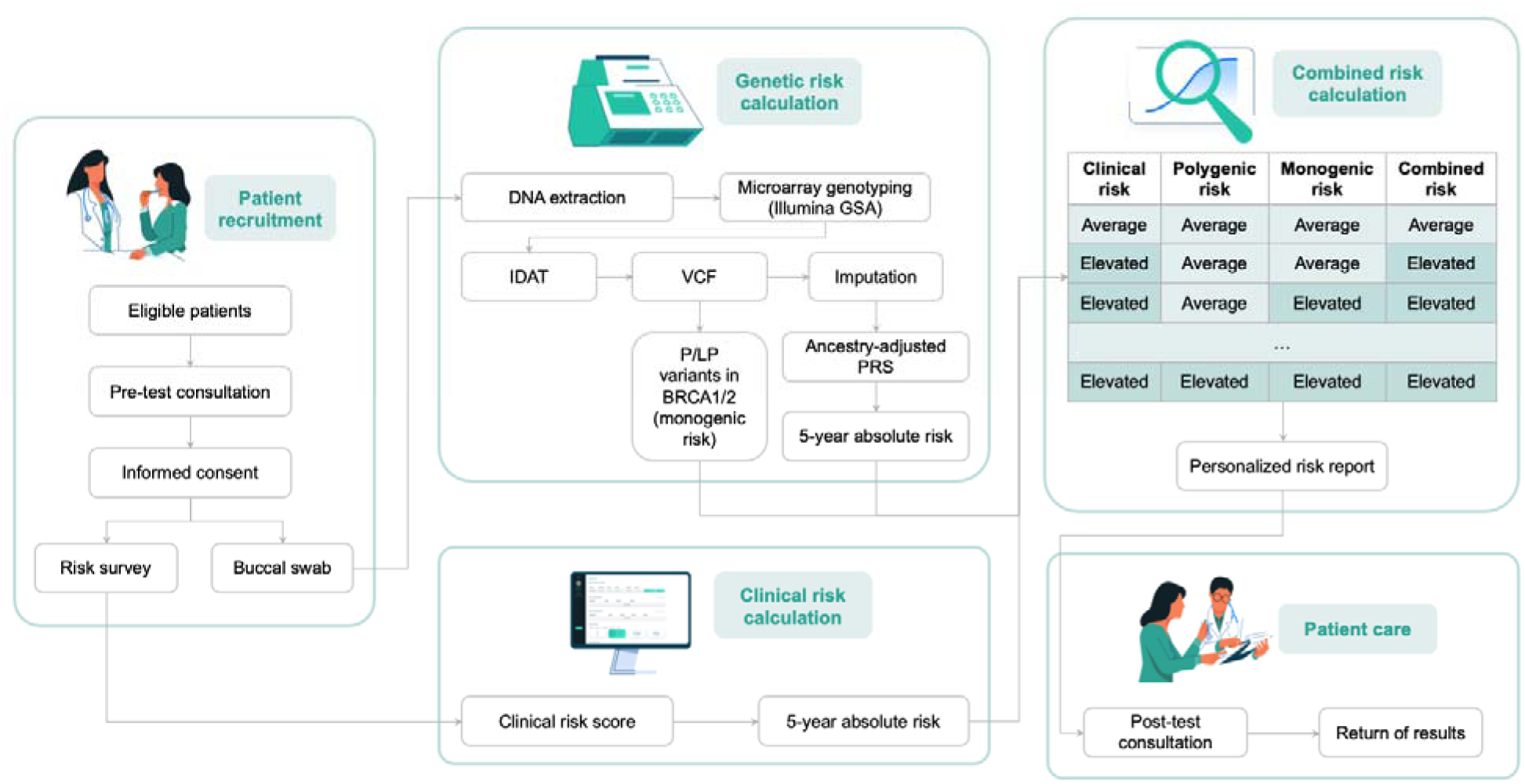
Personalized breast cancer risk assessment workflow. During the pre-test consultation, participants provide informed consent, complete a risk questionnaire, and submit a buccal sample. This sample undergoes array genotyping, followed by bioinformatic analysis to calculate ancestry-adjusted Polygenic Risk Scores (PRS) using the Mavaddat model and to identify known pathogenic variants in BRCA1/2. Concurrently, responses from the clinical risk survey are analyzed according to the Gail model. The single-factor risk results are then combined to determine a categorical risk outcome, which is communicated to participants in a personalized risk report during the post-test consultation.

To validate the accuracy of the aforementioned reporting workflow, we assembled complementary datasets for both pre-clinical and clinical validation. The pre-clinical validation study utilized well-characterized reference materials from the Genome In a Bottle (GIAB) project^18^, encompassing five samples tested in replicates, and Horizon reference standard cell lines^19^ with known mutations in BRCA1/2 genes, comprising two samples also tested in replicates. In addition, a mock dataset simulating responses to the clinical risk survey was used to confirm the correct implementation of the clinical risk model. The clinical validation study involved a case/control cohort, consisting of female breast cancer patients (cases) and healthy females (controls), recruited from MRCCC Siloam Hospitals Semanggi and other locations (**Figure 2**). Following participant triage and biological sample quality control, 305 individuals remained eligible for final analysis, comprising 149 cases and 156 controls (48.85% and 51.15% of the study population, respectively). The demographic breakdown revealed 114 participants of Indonesian Chinese ancestry and 191 from other Indonesian backgrounds (**Supplementary Table 1**), with mean ages of 47.86 years (± 8.20) for cases and 44.26 years (± 7.88) for controls.

**Figure 2:**
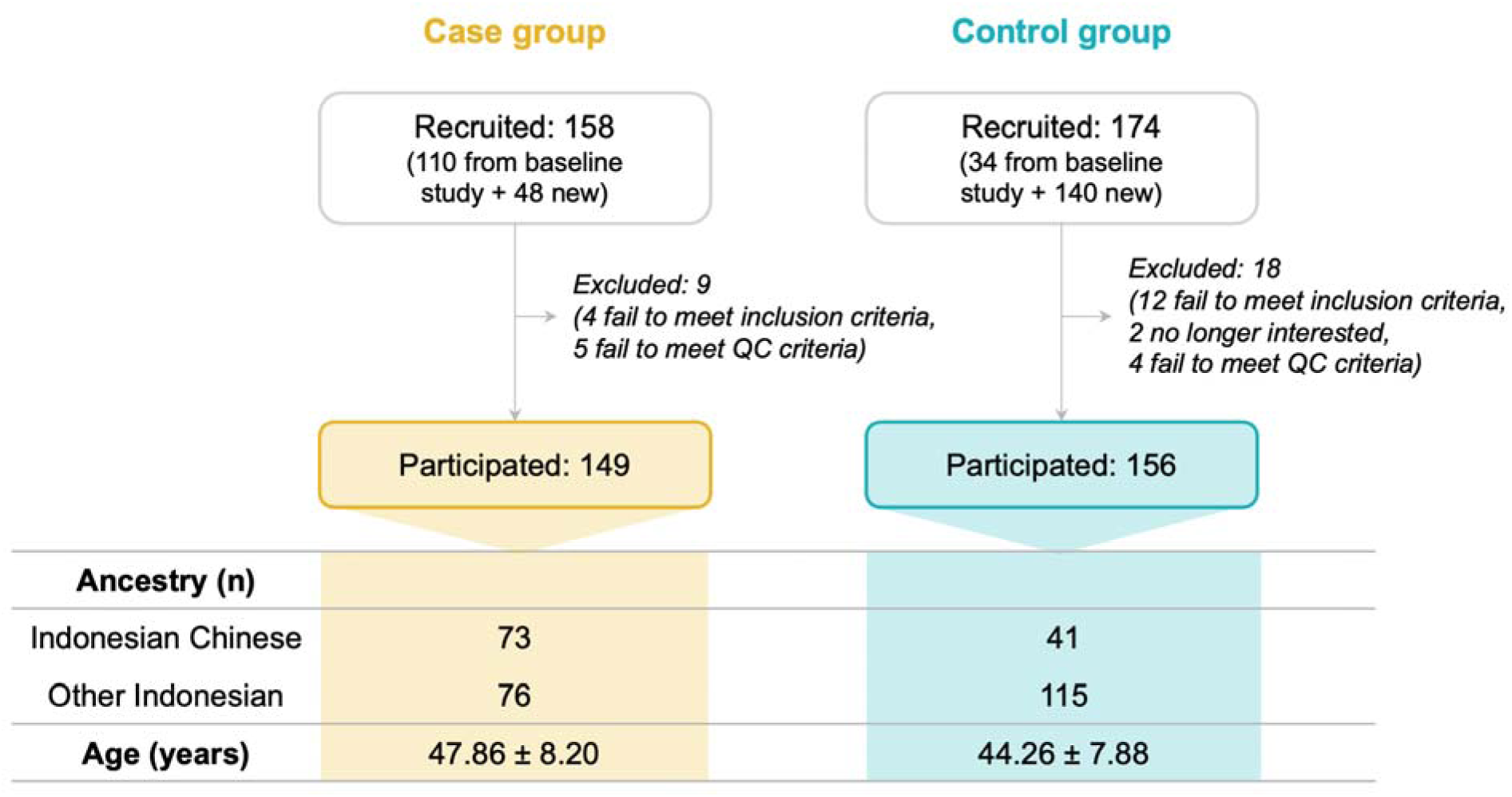
Participant recruitment and demographic breakdown in the clinical validation study. The clinical validation study employed a retrospective case/control cohort, consisting of female breast cancer patients (cases) and healthy females (controls), primarily recruited from MRCCC Siloam Hospitals Semanggi and other locations. Initially, 158 cases and 174 controls were enlisted. Following triage and biological sample quality control, 149 cases and 156 controls remained, totalling 305 eligible participants. The study maintained a balanced distribution with 48.85% of the participants in the case group and 51.15% in the control group. Demographic analysis revealed a diverse participant pool, encompassing 114 individuals of Indonesian Chinese descent and 191 from other Indonesian backgrounds. The mean ages were 47.86 years (± 8.20) for cases and 44.26 years (± 7.88) for controls.

### Accuracy of the Gail clinical risk model in the Indonesian population

Firstly, we evaluated whether the Gail model can be applied successfully to the local population. We first seeked to validate our tool by comparing answers from our data analysis pipeline to those from the NIH BRCAT tool^20^. We relied on simulated dataset with mock answers, which has been manually generated to cover a range of ethnicities and risk outcomes and utilized Pearson correlation analysis to compare the clinical risk scores from both methods (see Methods). We observed a very strong correlation (Pearson correlation 0.94, p-value 3.38×10^-15^) between the results of the NIH BRCA tool and the Bioinformatics pipeline used in our clinical workflow, supporting the validity of our software to calculate clinical risk.

Next, we evaluated the predictive accuracy of the Gail model by applying it to our patient cohort. We calculated 5-year absolute risk scores for all study participants and compared them between case and control groups. As expected, the case group exhibited higher scores (**Figure 3A**), with a mean of 0.76 (±0.44), compared to 0.54 (±0.31) for controls (p-value of 5.16×10^-04^). Notably, the difference in scores among groups persists when controlling for age and ethnicity as potential confounding factors in the study (p-value: 4.82×10^-02^). In addition, we observed an AUC of 0.67 with 95% CI [0.61,0.74] (**Figure 3B**), which aligns with previously reported outcomes for Western and Asian ancestry populations^15,21–25^. Altogether, these findings support the applicability of the Gail model to Indonesian populations.

**Figure 3:**
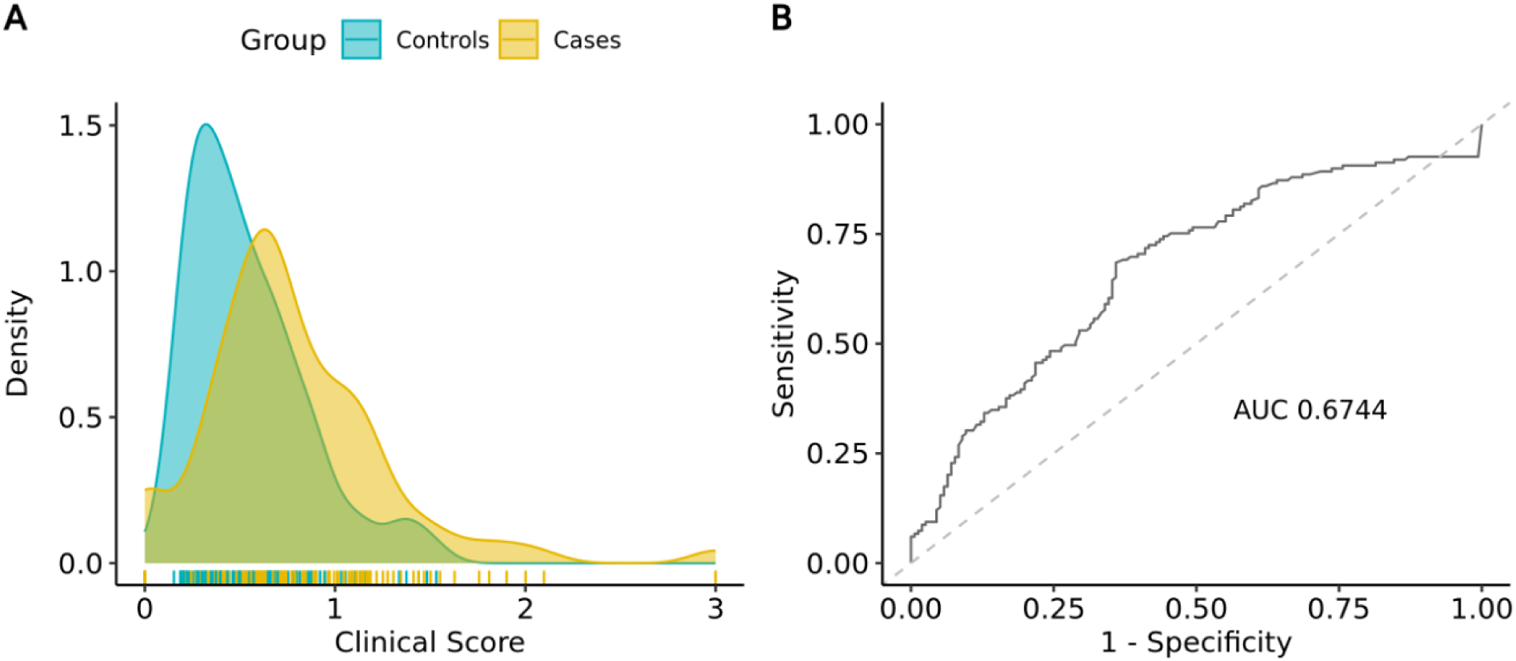
Distribution curve and predictive accuracy of the 5-year clinical risk score. (A) Distribution curve of the clinical risk score in cases vs. controls. Higher scores (0.76±0.44) are observed in cases compared to controls (0.54±0.31), with the difference being statistically significant (p-value: 5.16e-04). (B) ROC curve for the clinical risk score. The observed AUC is 0.674.

### Assessment of polygenic risk using the Mavaddat model

We next aimed to assess the genotyping accuracy of our array workflow by evaluating our ability to obtain correct genotype calls at PRS loci. For this assessment, we relied on a set of cell lines with well-established variant truthsets (Genome In A Bottle, GIAB)^18,26^, which we genotyped in-house. Assessing the accuracy of genotype calls in Mavaddat loci indicated high analytical sensitivity and specificity across all samples tested (99.25±0.46 and 96.89±0.50, respectively; **Table 1**). To further assess calling accuracy in each of the individual 313 loci of the PRS model, we expanded our evaluation dataset with 19 cell lines from the 1000 Genomes Project^27^, which combined with the previous GIAB dataset, covered 306 out of the 313 loci. This analysis showed that 98.69% of sites (302/306) had >95% concordance with the expected calls across all samples, thus demonstrating high per-site accuracy in our workflow (**Supplementary Figure 1**).

**Table 1:** Genotyping accuracy of Mavaddat PRS loci in GIAB samples (N=10).

| Metric |  | MEAN | SD | MIN | M<br>A<br>X |
| --- | --- | --- | --- | --- | --- |
| Callability | 95.37 | 2.29 | 93.29 | 99.68 |  |
| Genotype concordance | 97.81 | 0.33 | 97.38 | 98.18 |  |
| Analytical sensitivity | 99.25 | 0.46 | 98.66 | 100 |  |
| Analytical specificity | 96.89 | 0.5 | 96.3 | 97.6 |  |
| Precision | 99.58 | 0.57 | 99.64 | 100 |  |
| No-call rate | 3.86 | 1.91 | 0.32 | 6.07 |  |

Subsequently, we calculated PRS using PLINK, accounting for ancestry using established methods (see Methods). We compared ancestry-adjusted PRS distributions across cases and controls, and detected a significant difference across groups, with higher scores observed in the cases (0.63±0.97 vs. 0.24±0.88; p-value: 2.70×10^-04^), and an overall AUC of 0.63 (**Supplementary Figure 2**). Since our final report includes absolute risk scores instead of the relative risk reported by PRS, we also analyzed the distribution of 5-year absolute risk scores between groups. Notably, the trend previously observed for PRS persisted, with cases exhibiting higher risk scores compared to controls (0.94±0.48 vs. 0.68±0.38; p-value: 3.48×10^-07^, **Figure 4A**). Similar to previous observations for clinical risk, the difference among groups remained significant even after adjusting for age and ethnicity as potential confounding factors (p-value: 1.20×10^-03^). In addition, we observed an AUC of 0.67 with 95% CI [0.61,0.73] (**Figure 4B**), with is aligned with ranges previously reported in Western and Asian populations^16,28^, indicating that the Mavaddat model can be applied with equivalent predictive accuracy in the Indonesian population.

**Figure 4:**
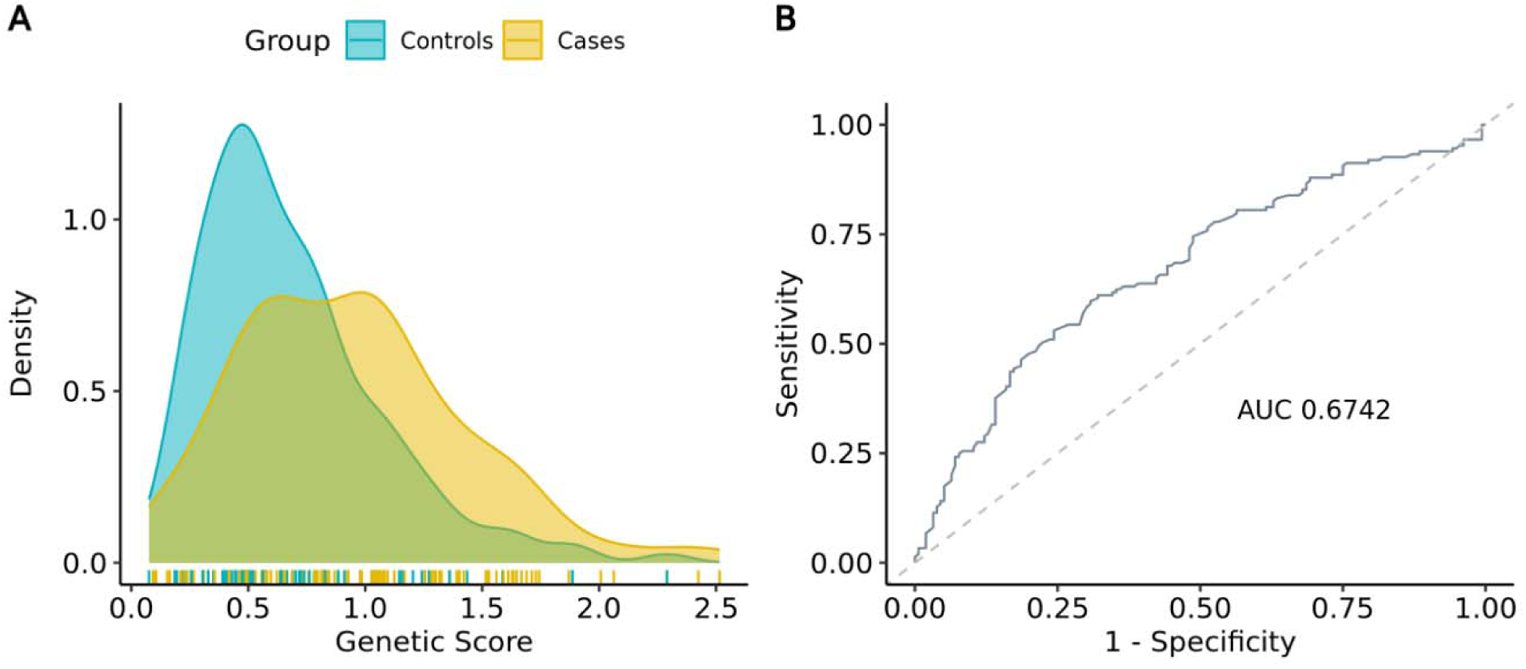
Distribution curve and predictive accuracy of the 5-year polygenic risk score. (A) Distribution curve of the genetic risk score in cases vs. controls. We observe higher scores in cases compared to controls (0.94±0.48 vs. 0.68±0.38; p-value: 1.28e-03). (B) ROC curve for the genetic risk score. The observed AUC is 0.6742.

Lastly, we sought to compare the genetic risk predictions from our software with those from an established third-party tool by analyzing array results from a subset of samples in our patient cohort (N=32) using both platforms (**Supplementary Table 2**). As expected, we noted a significant correlation in PRS results across both tools (Pearson correlation coefficient: 0.95; p-value: 6.11×10^-12^). However, we identified differences in the approaches each tool uses to interpret PRS into categorical risk outcomes. Our workflow translates PRS into a 5-year absolute risk, incorporating localized disease incidence and mortality rates, and utilizes a 1.7% threshold to distinguish between elevated and average risk. In contrast, the third-party tool categorizes PRS above the 91st percentile as high risk, equating to a ≥20% lifetime disease risk, and relies on a broader population reference to determine a patient’s percentile score. Notably, when comparing the predictive accuracies of the risk categories defined by each platform, we observed higher concordance with phenotypic outcomes in our software compared to the third-party tool (59.38% vs. 37.50%). This finding underscores the importance of considering localized factors when determining categorical risk outcomes from PRS.

### Identifying pathogenic mutations in BRCA1/2

The Illumina GSA chip, which we utilized to genotype samples in our study, encompasses a total of 5168 markers in the BRCA1 and BRCA2 genes, with 2900 of them being annotated as pathogenic/likely pathogenic mutations according to ClinVar^29^. Our analytical pipeline has been developed to report pathogenic mutations in these loci, prioritizing results from direct genotyping and preceding any imputation steps. Specifically, we focus on known pathogenic variants in ClinVar, with a confidence score of 2 or higher (see Methods).

In order to assess the performance of our workflow, we genotyped two Horizon reference material cell lines, HD793 and HD794, each engineered to contain mutations in BRCA1/2^30,31^. Each cell line was genotyped in triplicate, and we evaluated our results by comparing the obtained genotype calls to the verified mutations from Horizon, demonstrating 100% analytical sensitivity and specificity across 26 assessed markers (**Table 2**). After establishing the analytical validity of our workflow, we proceeded to interrogate the presence of pathogenic variants in our study cohort. We detected pathogenic mutations in 14 individuals, all of whom reassuringly belonged to the case group (**Supplementary Table 3**).

**Table 2:** Genotyping accuracy of verified BRCA1/2 mutations in Horizon reference materials (N=6).

| Metric | MEAN | SD | MIN | MAX |
| --- | --- | --- | --- | --- |
| Callability | 93.48 | 5.05 | 90.00 | 100.00 |
| Genotype concordance | 100.00 | 0.00 | 100.00 | 100.00 |
| Analytical sensitivity | 100.00 | 0.00 | 100.00 | 100.00 |
| Analytical specificity | 100.00 | 0.00 | 100.00 | 100.00 |
| Precision | 100.00 | 0.00 | 100.00 | 100.00 |
| No-call rate | 6.51 | 5.058 | 0 | 10.00 |

### Performance evaluation of the combined risk model

In the final stage of our data analysis workflow, we integrate various risk predictions - clinical, polygenic, and monogenic - to determine an overall risk category. Individual risk factors are initially converted into a categorical outcome as follows: 5-year absolute risk scores for clinical and polygenic risk are categorized as either elevated or average using a 1.7% cut-off, while monogenic risk is deemed elevated if a pathogenic variant is detected. Subsequently, a consolidated risk category is then established by selecting the highest risk classification from the three inputs (**Figure 1**).

To assess the accuracy of our combined risk predictions, we first evaluated the impact on accuracy when combining clinical and polygenic risk, as opposed to relying on single-risk factors alone. The analysis yielded an AUC of 0.70 [95% CI: 0.64,0.76] for the two-factor combined risk model, higher than that of single-factor risk models (**Figure 5A**). Next, we examined the proportion of study participants classified as either average or elevated risk within the case and control groups, taking into account the three-factor consolidated risk categories. Our analysis revealed a higher proportion of elevated risk predictions in the case group compared to the controls (15% vs. 4%, respectively; **Figure 5B**), a trend that was statistically significant with an odds ratio of 3.81 and a p-value of 1.3×10^-03^. Lastly, we estimated the Positive Predictive Value (PPV) and Negative Predictive Value (NPV) of our assay, with values of 78.13% and 54.58%, respectively.

**Figure 5:**
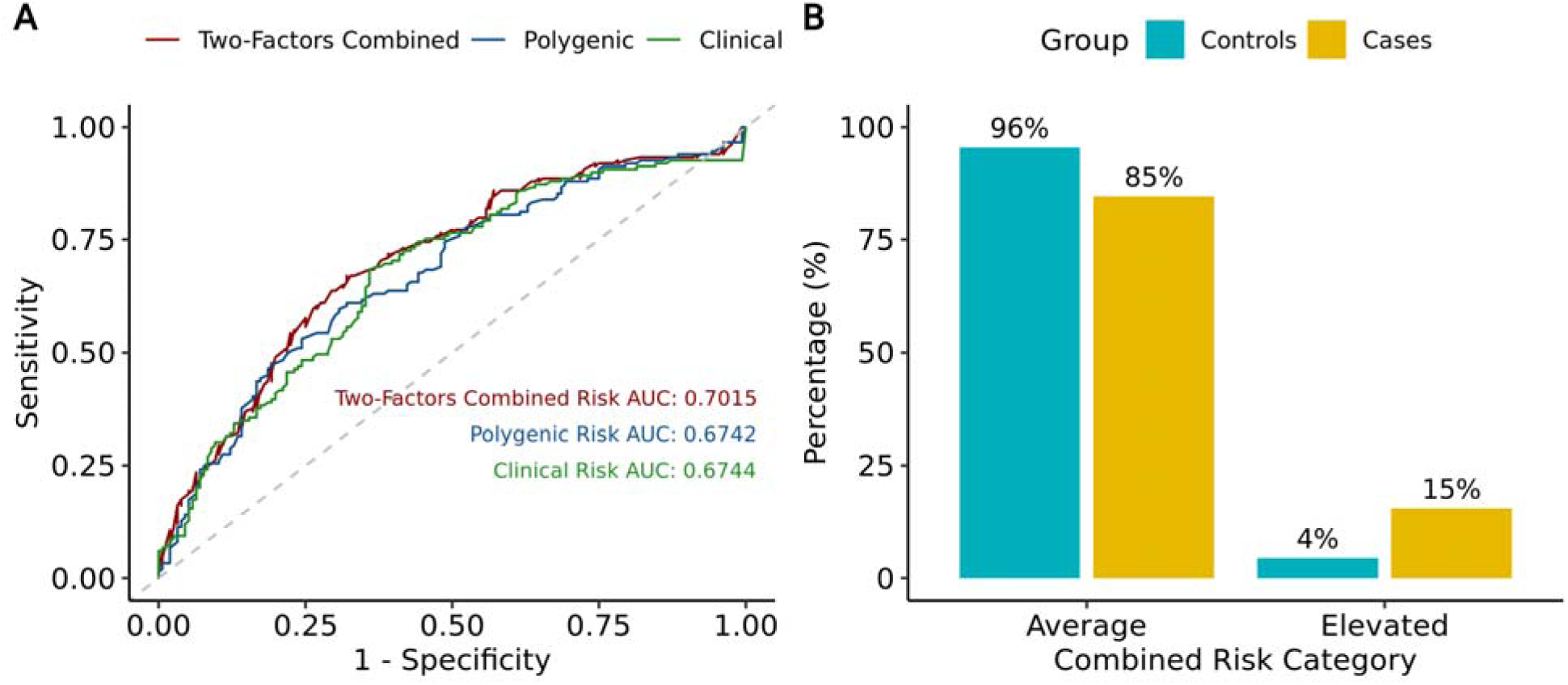
Combined risk outcomes and comparison of AUCs. (A) Comparison of ROC curves between clinical, polygenic and two-factor combined risk (clinical and polygenic) (B) Percentage of samples categorised as average or elevated based on three-factor combined risk (clinical, polygenic and monogenic).

## Discussion

Early detection of breast cancer significantly enhances patient prognosis and reduces mortality rates, yet current screening strategies predominantly adhere to a one-size-fits-all approach. Notably, not all countries have established population screening programs to promote mammography screening, and even in countries that have, the uptake rate remains low. For instance, in Indonesia, breast cancer screening has not yet been integrated into the national health agenda. Meanwhile, in Singapore, where it has been established, only a mere 28% of primary educated women undergo screening, and 30% of diagnoses occur in women below the suggested screening age^32,33^. Emerging risk-based screening approaches promise better patient stratification and increased screening rates. However, the prevalent ancestry bias in the development of underlying risk models poses a significant challenge for their widespread adoption, particularly for PRS models, which rely on common genetic variants that can be markedly influenced by population-specific allele frequencies. While creating new models based on diverse ancestries is technically feasible, the scarcity of data presents a substantial barrier, making the evaluation of the applicability of existing models to various ancestries a critical task. In this context, our study aims to assess the clinical validity of two established risk prediction models, the Mavaddat and Gail models, for the Indonesian population, which we combine with monogenic risk factors, particularly pathogenic mutations in BRCA1/2 genes. In addition, we propose a personalized breast cancer risk assessment workflow based on array genotyping as a first step towards the implementation of risk-based screening in the region.

To validate the predictive accuracy of the Gail and Mavaddat models in the Indonesian context, we conducted a case-control study involving 305 participants of local descent. Our analysis revealed that the observed AUC of 0.67 for the Gail model aligns with the range documented in published studies^15,21–25^. For genetic risk, we observed an AUC of 0.63 for ancestry-adjusted PRS, indicating a slight regression in performance compared to the original study (AUC=0.64), which utilized samples from the UK Biobank^16^. The AUC, a metric derived from the Receiver Operating Characteristic (ROC) curve, is commonly used when evaluating the predictive accuracy of a model, and a higher AUC implies that the model has a higher true positive rate and a lower false positive rate, which is crucial in minimizing misclassifications in risk prediction. Altogether, our observations on the AUC of ancestry-adjusted PRS suggest the potential for developing more accurate models in the Asian population, a prospect that could become a reality as larger Asian cohorts become available to identify population-specific variants. Current population-genomic initiatives, such as the Biomedical and Genome Science Initiative (BGSi) in Indonesia, are poised to bridge this gap. It is also important to emphasize that our method of interpreting risk scores into categories relies on absolute risk, not relative risk. By incorporating localized data on disease incidence and mortality from Indonesian statistics, we have achieved improved performance in risk scores compared to solely interpreting PRS (AUC of 0.67 for absolute risk scores vs. 0.63 for ancestry-adjusted PRS). This localized approach demonstrates a more significant impact when comparing our risk prediction results with those from a third-party software that relies on a broader population reference to derive risk categories based on PRS percentiles, as demonstrated by the higher concordance with phenotypic outcomes in our software compared to the third-party tool (59.38% vs. 37.50%). Altogether, our observations show, for the first time, the applicability of the Gail and Mavaddat models in the Indonesian context and highlight the potential for further enhancing performance through the creation of new models based on localized datasets.

While the results presented above showcase the performance of both clinical and PRS models individually, it underscores that the best accuracy is achieved when these models are combined (AUC of 0.70 for the combined risk model vs. 0.67 for single-factor models). This finding aligns with previous research, suggesting that a multi-faceted approach to risk prediction surpasses the efficacy of single-factor models. For example, research conducted by Hurson et al^34^ utilizing multiple datasets of European ancestry illustrated that integrating classical factors with PRS can enhance the detection of individuals at increased risk. Yang et al reported a comparable trend in a dataset encompassing various Asian populations, including Chinese, Japanese, Korean, Thai, Singaporean, and Malaysian groups^35^. While the integration of various risk factors can enhance predictive capabilities, it also introduces complexity into model development, validation, and interpretation. Our research addresses these challenges and extends previous observations to include data from the Indonesian population, thereby contributing additional demographic variability into the analysis of risk factors across heterogeneous populations. In addition, emerging studies suggest that developing risk models with additional input data can substantially enhance predictive accuracy. In this context, the emergence of advanced models such as BRISK (Allman et al., 2023), which aggregates mammographic density, polygenic risk, and clinical factors, and BOADICEA^36^, which incorporates a comprehensive range of inputs including detailed family history, genetic data, PRS, and lifestyle factors. Thus, there is potential to further enhance the accuracy of models in the Indonesian population through the inclusion of additional datasets.

In addition to evaluating AUCs for risk models, our study also includes an assessment of the genotyping accuracy of our risk prediction workflow. To this end, we utilized GIAB reference materials and focused the performance evaluation on the markers required for calculating PRS under the Mavaddat model. We determined the analytical sensitivity and specificity of our array genotyping workflow to be 99.25% (±0.46) and 96.89% (±0.5), respectively. Such evaluation is vital in establishing a clinical-grade testing workflow, as it ensures the accuracy of results inputted into the risk calculation software. Most importantly, it should be conducted with the target loci in mind. Given that PRS models encapsulate common variants from numerous genomic loci, which can vary significantly in number and nature between models and considering that different models might encompass a larger representation of SNPs or INDELs, genome-wide evaluation alone may not necessarily be representative of the performance at selected loci. Additionally, we assessed genotyping accuracy in BRCA1/2 genes using Horizon reference materials, emphasizing that such assessments, while challenging to conduct, should be pursued when possible.

In conclusion, our study underscores the efficacy of the Gail and Mavaddat models in predicting breast cancer risk in the Indonesian population, demonstrating a performance comparable to studies conducted in other demographics. Furthermore, we illustrate that a combined risk model, which integrates clinical, polygenic and monogenic risk scores, excels in accuracy compared to single-factor models. The study serves as a preliminary yet promising exploration, as larger sample sizes will be required to fully capture the rich genetic diversity in Indonesia. The emergence of novel datasets and biobanks from population genomics initiatives like BGSi promises to facilitate further evaluation of risk models, and potentially foster the development of new models finely tuned to local populations. Beyond accuracy evaluations, the integration of personalized risk assessment into routine clinical practice requires a broader focus that also encompasses considerations of utility and cost-efficiency. While the practical application of enhanced breast cancer risk assessment models in hospital settings remains largely unexplored, pilot studies in other health domains show evidence that it could potentially foster behavioral changes, promoting risk-reducing behaviors among high-risk individuals^37^. This could in turn significantly reduce the incidence of advanced-stage breast cancer cases and the associated healthcare costs^12,36,38^. Moreover, this type of personalized risk assessment could be especially beneficial in regions such as Indonesia, where population-based screening is unavailable and where, given the population size, resources are limited, aiding in the identification of individuals who would benefit most from more frequent monitoring. Current global initiatives, such as “Our Future Health” in the UK^39^ and the Genomes2Veterans study in the US^40^, are leading the way in integrating personalized risk assessment in healthcare settings, thereby setting a precedent for further localized studies to emulate. Although still in its infancy, our collective efforts, in conjunction with others, mark the onset of a transformative era in risk assessment, steering towards a more personalized and proactive approach to healthcare.

## Methods

### An end-to-end workflow for personalized breast cancer risk assessment

As part of this study, we developed an end-to-end workflow for personalized breast cancer risk assessment. A visual representation of the workflow is provided in **Figure 1**, with details on the methods described below.

#### Pre-test consultation

Eligible female participants (women aged 25-75 who have never been diagnosed with breast cancer and have no known mutations in BRCA1 or BRCA2) initially undergo a pre-genetic consultation, either on-site or online, where they receive information about the test. After the consultation, they are asked to consent to the collection of buccal samples for genetic testing and to complete a questionnaire that gathers non-genetic breast cancer risk factors, as outlined in the Gail model (**Supplementary File 1**). Buccal samples are then collected using OraCollect (cat. no. DNA OCR-100, provided by DNA Genotek).

#### gDNA Extraction and genotyping

Genomic DNA (gDNA) is extracted using the Monarch® Genomic DNA Purification Kit (cat. no. T3010 from NEB). The extraction procedure adheres to the manufacturer’s instructions, incorporating an additional dry-spin step at maximum speed for 1 minute following the second buffer washing step. The quality and concentration of the gDNA extracts are measured using BioDrop-µLITE. The acceptance criteria for DNA quality adhere to the manufacturer’s guidelines for the extraction kit, specifically requiring absorbance ratios of A260/230 and A260/280 to be greater than 1.7, and a DNA yield exceeding 500 ng.

#### Array Genotyping

Genotyping is conducted using standard processing on the Illumina GSA chip (Infinium Global Screening Array-24 Kit) by Genomic Solidaritas Indonesia. Raw data files (IDAT files) are converted to VCF format using iiap-cli & GTCtoVCF Illumina software (genome build GRCh37). Missing calls are inferred by performing imputation with Eagle2^41^ and minimac4^42^, using the 1000 Genomes project as the reference panel.

#### Calculation of clinical risk

The calculation of clinical risk is based on the Gail model^13^. Five-year absolute risk scores are determined using responses from the pre-test consultation questionnaire, which includes information on the patient’s age, age at menarche and first full-term pregnancy, number of first- degree relatives with breast cancer, history of breast biopsy, presence of breast biopsy with atypical hyperplasia, and ethnicity.

#### Calculation of polygenic risk

Genetic risk is assessed using the Mavaddat PRS model^16^. Initially, microarray results are subsetted to focus on the 313 markers specified in the model. Direct genotyping and imputation results are integrated, prioritizing microarray data, and supplementing with imputation results only when the INFO SCORE exceeds 0.8.

Subsequently, genotype calls are utilized for PRS calculation through PLINK^43^. Briefly, this process involves computing a weighted average of alleles present in each individual’s genetic profile across the 313 variants specified in the model. Missing variants are inferred using the --read-freq option and allele frequency data from the GNOMAD database^44^. Next, the raw PRS scores from PLINK are standardized as Z-scores, using the mean and standard deviation from a Southeast Asian cohort (MEC study^11^). These standardized scores are then adjusted for population structure according to methods outlined by Hao et al., 2022, employing a linear regression model that incorporates the first four principal components derived from the 1000 Genomes dataset^27^. Ultimately, the ancestry-adjusted PRS are translated into 5-year absolute scores using established methods^9^, which factor in incidence and mortality data pertinent to the Indonesian population^28^.

#### Calculation of monogenic risk

The BRCA1 and BRCA2 genes were selected as the target for monogenic risk calculations. All detected variants in these (based on direct genotyping from the Illumina GSA chip) genes were annotated using Nirvana 3.18.1^45^. Following the annotation process, additional filtering was performed according to the following criteria: (i) the variant’s significance is either pathogenic or likely pathogenic; (ii) it possesses a ClinVar review rating of >=2 stars; (iii) there is no conflicting interpretation; (iv) it is a non-reference variant; and (v) it has a genotype quality score of >=3. The determination of the monogenic risk category is based on whether a pathogenic or likely pathogenic variant is detected in the sample.

#### Calculation of combined risk

Initially, individual risk factors are transformed into categorical outcomes in the following manner: 5-year absolute risk scores for clinical and polygenic risk are classified as either elevated or average, utilizing a 1.7% threshold, while monogenic risk is considered elevated if a pathogenic variant is identified. Following this, a unified risk category is determined by choosing the most elevated risk classification from the three inputs.

#### Post-test consultation

The risk assessment results are compiled into a personalized report that includes clinical and genetic risk scores and categories, combined risk categories, and customized health modification recommendations based on the test findings. Examples of these reports for both average and elevated risk individuals can be found in **Supplementary Files 2** and **3**. These risk reports are then presented to patients during a post-test consultation with an oncologist.

### Workflow validation

To validate the accuracy of our risk prediction workflow, we conducted both pre-clinical and clinical validation studies. On the one hand, the pre-clinical validation study employed well-characterized reference materials from the Genome In a Bottle (GIAB) project, along with a mock dataset simulating responses to the clinical risk survey, to assess our ability to accurately genotype variants of interest and verify the correct implementation of the Gail model. On the other hand, clinical validation, executed as a case-control study, aimed to evaluate the predictive accuracy of the selected models (including both the clinical risk and PRS, as well as their combined risk assessment) specifically within the Indonesian population.

#### Pre-clinical validation

##### Confirming implementation of the Gail model

A pre-clinical validation study was conducted to assess the accuracy of our clinical risk prediction algorithm, which was developed based on the Gail model. This validation employed a mock dataset created without any ties to actual patients or individuals. The dataset consisted of manually generated clinical survey responses, encompassing a wide range of ethnicities, clinical risk factors, and risk outcomes. Using this mock dataset as input, we proceeded to calculate a clinical risk score using our algorithm. Simultaneously, the mock dataset was also analyzed using the NIH Breast Cancer Risk Assessment Tool (BRCAT) to determine the anticipated clinical risk score. Upon completing both analyses, a comparison was undertaken using Pearson correlation analysis.

##### Assessing genotyping accuracy in Mavaddat PRS markers

A separate validation study was conducted to assess the performance of our genotyping algorithm. The validation process employed Genome in a Bottle (GIAB) samples (HG001-005), with each being genotyped in duplicate. Each sample underwent processing through our data analysis pipeline to derive genotype calls at each of the 313 target sites. Resulting VCF files were compared against their respective truth sets. The accuracy of the genotyping was assessed on a per-sample basis by calculating various metrics, including:

- Callability: The percentage of loci successfully genotyped out of the 313 loci in the PRS model.
- Genotype concordance: The percentage of genotyped sites with a correct call.
- Analytical sensitivity: The percentage of variant sites correctly identified.
- Analytical specificity: The percentage of non-variant sites correctly identified.
- Precision: The percentage of variants correctly genotyped relative to the number of reported variants.

Additionally, we genotyped 18 additional 1KGP cell lines to facilitate a per-site assessment of variant calling accuracy. The truthset for this dataset consisted of 30X WGS results from the Registry of Open Data in AWS^27^. The combined set of results, including 5 GIAB and 19 1KGP cell lines, covered 306 out of the 313 PRS loci. Each site was evaluated for concordance against the truthset across all samples and deemed concordant if the results were correct in more than 95% of the samples.

##### Comparative analysis of PRS against an established method

We validated the implementation of the Mavaddat PRS model in our algorithm through a comparative analysis with a genetic risk prediction tool from a third-party software company. We utilized a subset of the samples from the current study, which included 12 healthy individuals and 20 breast cancer patients. The evaluation was based on two predefined criteria: first, we measured the correlation between the polygenic risk scores; and second, we assessed the overlap in categorical outcomes with the phenotypes present in our cohort.

##### Assessing genotyping accuracy in BRCA1/2 genes

To assess genotyping accuracy in BRCA1 and BRCA2 genes, we utilized two distinct reference materials from Horizon (HD793 and HD794), each engineered to contain mutations in these genes, and evaluated a total of 26 genomic loci. Each sample was genotyped three times and processed through our data analysis pipeline to derive genotype calls at each target site. The resulting VCF files were compared against Horizon’s verified mutations, and genotyping accuracy was assessed using the performance metrics introduced above (see “Assessing genotyping accuracy in Mavaddat PRS markers”).

#### Clinical validation study

##### Study design and ethical approval

To assess the accuracy of our risk prediction workflow in a real-world setting, we initiated a case/control study (TRIP), during which healthy participants and breast cancer patients were recruited over a span of two years (from May 2021 to February 2023). This study received ethical approval from the MRCCC Siloam Hospitals IRB, under the approval number 005/EA/KEPKKRSMRCCC/X/2020. Amendments to the study protocol, including the inclusion of a larger sample size, have also been approved by the IRB, under the approval number 005/EA/KEPKKRSMRCCC/X/2022.

##### Sample size calculation

Power analysis was conducted to determine sample size of the study using Python’s statsmodel module^46^. We estimated the sample size with power analysis based on t-test for two independent groups. The effect size is calculated using Cohen’s D formula:

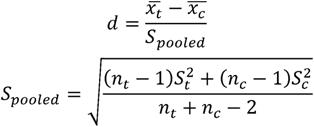

where *d* is effect size, *x̄* is the mean of the PRS, *S* is the standard deviation, *n* is the number of samples, and the subscript *t* and *c* refers to the treatment and control groups respectively. The mean and standard deviation are based on literature^33^. This calculation provided us with an effect size of 0.396. The estimated sample size for two-tailed test is calculated using the following formula:

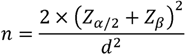

where *n* is the sample size for each group, *Z_α_*_/2_ is the critical value of *α* and *Z_β_* is the critical value of *β*. Based on this analysis, we estimated that the minimum sample size would be 101 samples for both case and control groups, or a total of 202 samples, assuming a power of 0.8. We recruited additional samples to minimize error and anticipate dropouts.

##### Patient recruitment and enrollment

The case/control study included both breast cancer patients and healthy individuals, provided they met the established eligibility criteria (see **Supplementary Table 4**). Breast cancer patients were assigned to the case group, while healthy participants were categorized into the control group. Individuals not affiliated with Indonesian ethnic groups or of Chinese-Indonesian descent were excluded from the study to align with the research’s specific population focus. Additionally, participants who did not provide informed consent for participation and subsequent follow-up were also excluded from the study cohort.

Following the initiation of the recruitment process, we enlisted 191 female participants from MRCCC Siloam Hospitals Semanggi, both onsite and through online sessions via Zoom, to partake in the breast cancer risk prediction study. An additional 141 participants were incorporated from a baseline study, with consent for the utilization of their remaining DNA for further studies concerning breast cancer. Out of 332 participants, there were 158 (47.59%) cases and 174 (52.41%) controls. This dataset was later narrowed down to 314 participants after excluding 18 (5.42%) due to loss to follow-up (n=1), withdrawal (n=1), and failure to meet the inclusion criteria (n=16). This number was further revised to 305 participants after identifying samples that did not meet the quality control criteria for analysis (**Figure 2**).

The final cohort comprised a total of 305 female participants, including 149 individuals (48.85%) diagnosed with breast cancer (cases) and 156 individuals (51.15%) without the condition (controls). Notably, the demographic distribution ultimately included 114 participants of Chinese lineage, consisting of 73 cases and 41 controls, as well as 191 participants of Indonesian heritage, encompassing 76 cases and 115 controls.

#### Statistical analysis

##### Comparing PRS and risk score distributions

An independent samples t-test was conducted to compare the clinical score, adjusted PRS, and genetic score between cases and controls. This test was utilized to determine whether significant differences existed between the two groups. Additionally, logistic regression analysis was performed to examine the relationship between the categorical outcome variables and the variables of interest, while controlling for the effects of age and ethnicity.

##### Calculation of area under the curve (AUC)

Receiver Operating Characteristics (ROC) analysis was conducted to assess the predictive performance of the clinical score, adjusted PRS, genetic score, and combined risk. The ROC curve, generated using the R package “pROC”^47^, provided the area under the curve (AUC) which was used to quantify the overall discriminative ability of our models. To calculate the AUC for combined risk, we first listed a set of possible thresholds using pROC. For each threshold value, we applied the threshold to both the clinical and genetic scores to infer a categorical combined risk. We then calculated the sensitivity and specificity of each threshold. Finally, we plotted the ROC curve and calculated the AUC based on the sensitivity and specificity values.

##### Calculation of odds ratio (OR)

To assess the strength of the association between the dependent variable (cases and controls) and combined risk categories (elevated and average), a contingency table was created using the “epitools” package^48^. The odds ratio was then calculated based on the contingency table using the “oddsratio” function, providing a measure of the association strength between the variables of interest.

## Supplementary Files

**Supplementary File 1. Clinical risk questionnaire.** This questionnaire is administered to study participants after obtaining their informed consent.

**Supplementary File 2. Example of personalized risk report for an individual with average risk.** Patients will be provided with a comprehensive report delineating their average risk of developing breast cancer. This report includes an analysis of both genetic and clinical risk factors and offers guidance on steps that can be taken after the risk assessment.

**Supplementary File 3. Example of a personalized risk report for an individual with elevated risk.** Similar to Supplementary File 2, the following report is customized for individuals exhibiting an elevated risk. The interpretation of this report is to be conducted under the guidance of a medical professional.

**Disclaimer\***

Images of individuals included in the example report are models specifically incorporated as part of the report design for illustrative purposes. Furthermore, we emphasize that all patient information is entirely fabricated for similar reasons.

## Data Availability

All data produced in the present study are available upon reasonable request to the authors.

## Acknowledgements

We would like to thank dr. Dismas A. Chaspuri, SpB of MRCCC Siloam Hospitals Semanggi for his assistance in recruitment by referring his patients to participate in this study. We would also thank Ns Ria and Ns Surtati for their assistance during the recruitment process.

---

**Supplementary Figure 1:**
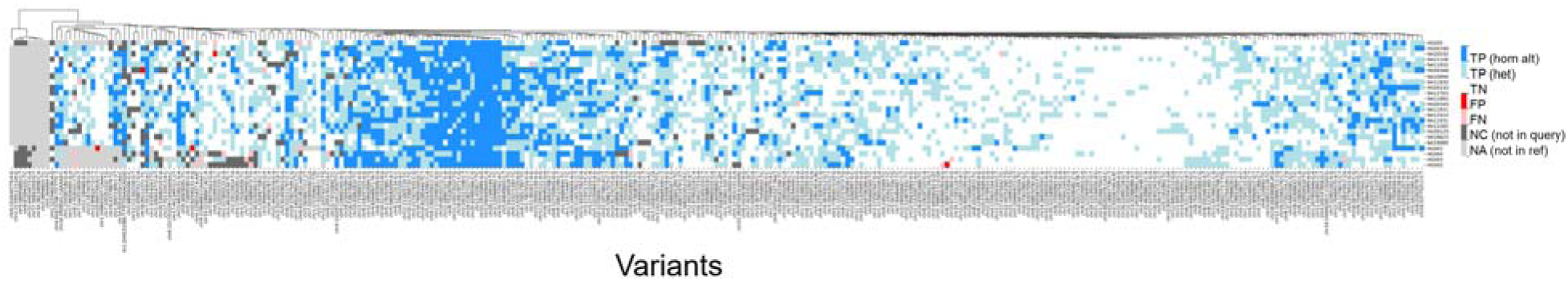
Per-site calling assessment of Mavaddat PRS loci. Heatmap depicting the genotyping concordance for 313 variants of interest associated with breast cancer risk prediction out of 23 samples. Concordance was assessed by comparison to reference calls based on whole genome sequencing of 1KGP or Genome-In-A-Bottle samples. TP (blue): True positive homozygous alternate call, TP (light blue): True positive heterozygous call, TN (white): true negative, FP (red): false positive, FN (pink): false negative, NC (dark grey): No call in the query sample, NA (light grey): No call in the truth set.

**Supplementary Figure 2:**
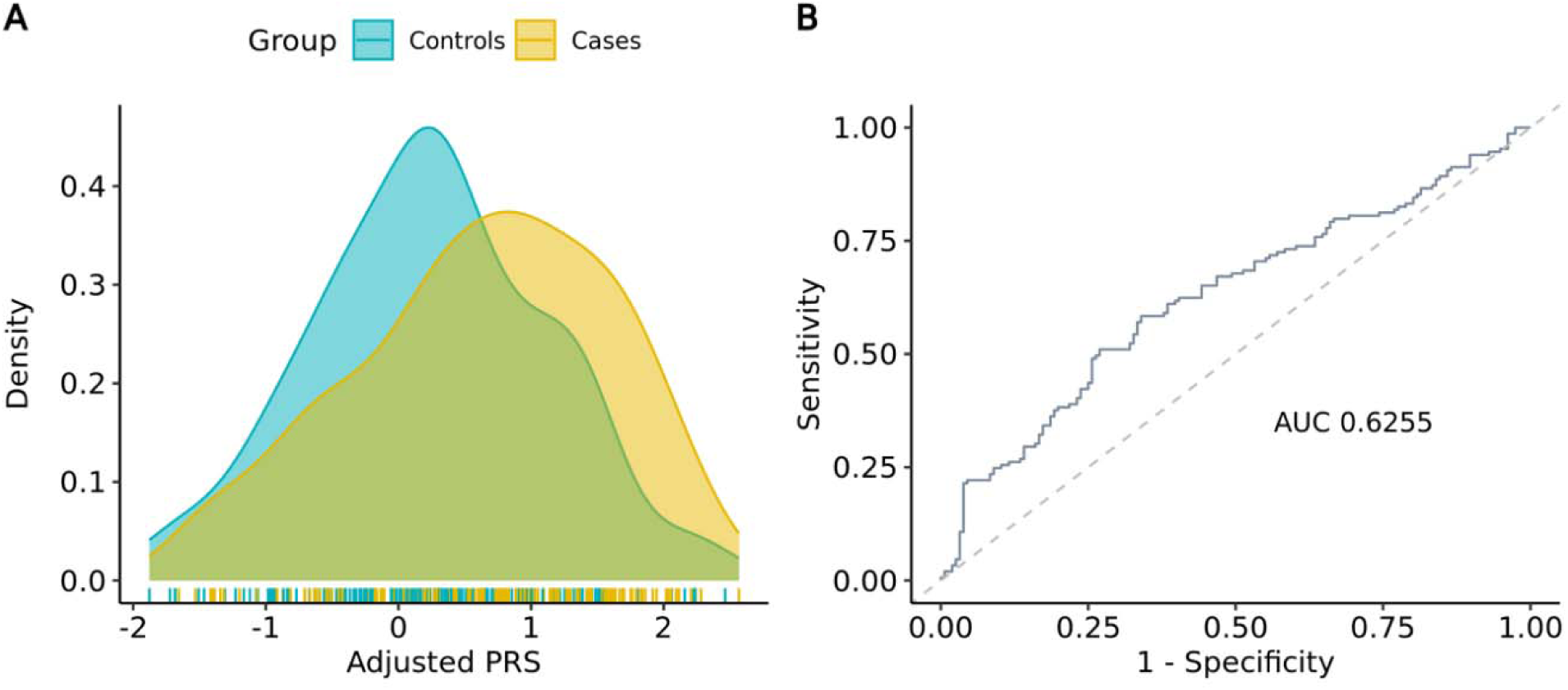
Distribution curve and predictive accuracy of ancestry-adjusted PRS. (A) Distribution curve of adjusted PRS in cases vs. controls. Higher PRS were observed in the case group compared to the controls (0.63±0.97 vs. 0.24±0.88; p-value: 2.20e-03). (B) ROC curve for adjusted PRS. The observed AUC is 0.6255.

**Supplementary Table 1:** Detailed ancestry breakdown for participants in the clinical validation study.

| **Ethnicity** | **Case (n, %)** | **Control (n, %)** |
| --- | --- | --- |
| Chinese | 73 | 41 |
| Indonesian | 75 | 116 |
| Jawa | 35 | 35 |
| Sunda | 6 | 14 |
| Batak | 6 | 14 |
| Bugis | 0 | 6 |
| Flores | 0 | 6 |
| Bali | 0 | 4 |
| Minangkabau | 3 | 1 |
| Manado | 3 | 2 |
| Betawi | 3 | 1 |
| Ambon | 0 | 3 |
| Maluku | 1 | 0 |
| Padang | 1 | 2 |
| Tolaki | 1 | 0 |
| Melayu | 1 | 0 |
| Timor | 1 | 0 |
| Nias | 1 | 0 |
| Aceh | 0 | 2 |
| Kalimantan Tengah | 0 | 1 |
| Other Sumatera* | 2 | 4 |
| Madura | 0 | 1 |
| Multi ethnicity** | 9 | 16 |
| Others*** | 3 | 3 |

\*Other Sumatera includes Palembang (1), Lampung (1), Komering Ulu (1), and others
\*\*Multi ethnicity refers to participants with more than one ethnicity, which includes Jawa-Sunda (6), Betawi-Sunda (2), Jawa-Betawi (2), etc.
\*\*\*Others refers to participants who did not specify the specific Indonesian ethnicity they derived from

**Supplementary Table 2:** Comparative analysis of genetic risk predictions with a third-party tool.

| Sample ID | Phenotype | PRS in-house | Genetic risk category in-house | PRS third-party tool | Genetic risk category third-party tool |
| --- | --- | --- | --- | --- | --- |
| 41001901220129_BC | CANCER | 1.233911504 | AVERAGE | 1.328450579 | AVERAGE |
| 41001901220150_BC | CANCER | 2.643061947 | ELEVATED | 2.315536285 | ELEVATED |
| 41001901220153_BC | CANCER | 1.880442478 | ELEVATED | 1.261834573 | AVERAGE |
| 41001901222079_BC | CANCER | 2.305097345 | ELEVATED | 2.205859718 | ELEVATED |
| 41001901222094_BC | CANCER | 2.306477876 | ELEVATED | 0.986018963 | AVERAGE |
| 41001901222177_BC | CANCER | -1.438454867 | AVERAGE | -1.733377778 | AVERAGE |
| 41001901222179_BC | CANCER | 2.14980531 | ELEVATED | -0.465485248 | AVERAGE |
| 41001901222199_BC | CANCER | 1.029750442 | AVERAGE | 0.308784296 | AVERAGE |
| 41001901222200_BC | CANCER | 2.31539823 | ELEVATED | -0.469953448 | AVERAGE |
| 41001901222272_bc | CANCER | 1.513437168 | AVERAGE | 1.308739488 | AVERAGE |
| 41001901222278_BC | CANCER | 0.637667257 | AVERAGE | 0.693980954 | AVERAGE |
| 41001901222285_BC | CANCER | 0.423437168 | AVERAGE | 0.251828443 | AVERAGE |
| 41001901222296_BC | CANCER | 1.803911504 | AVERAGE | 1.885205009 | ELEVATED |
| 41001901222389_BC | CANCER | 2.103327434 | ELEVATED | -0.305676435 | AVERAGE |
| 41210853008755_BC | NORMAL | 1.431745133 | AVERAGE | 0.917035367 | AVERAGE |
| 41210853008783_BC | NORMAL | 2.140548673 | ELEVATED | 1.881677965 | ELEVATED |
| 41210853015045_BC | CANCER | 1.654389381 | AVERAGE | 0.9732685 | AVERAGE |
| 41210853015093_BC | NORMAL | 0.177699115 | AVERAGE | -0.478522053 | AVERAGE |
| 41210853015147_BC | NORMAL | 0.492143363 | AVERAGE | 0.581812225 | AVERAGE |
| 41210853016184_BC | NORMAL | 2.28780531 | ELEVATED | 1.906911005 | ELEVATED |
| 41210853016186_BC | CANCER | 1.83920354 | ELEVATED | 1.424638125 | AVERAGE |
| 41210853016189_BC | CANCER | -0.361402655 | AVERAGE | -0.726921715 | AVERAGE |
| 41210853016254_BC | CANCER | 2.133504425 | ELEVATED | 1.579791975 | ELEVATED |
| 41210853016290_bc | NORMAL | -0.731976991 | AVERAGE | -0.628195591 | AVERAGE |
| 41210853016297_BC | NORMAL | 2.261840708 | AVERAGE | 1.888042727 | ELEVATED |
| 41210853016318_BC | NORMAL | -0.211773982 | AVERAGE | -1.888782596 | AVERAGE |
| 41210853016330_BC | NORMAL | 1.495254867 | AVERAGE | 1.25358441 | AVERAGE |
| 41210853016336_bc | NORMAL | -0.290918053 | AVERAGE | -1.279861282 | AVERAGE |
| 41210853016343_BC | NORMAL | 2.309681416 | ELEVATED | 2.371723913 | ELEVATED |
| 41210853018000_BC | CANCER | 2.188743363 | ELEVATED | 1.054420291 | AVERAGE |
| 41210853018727_BC | CANCER | 1.464444248 | AVERAGE | 0.873173875 | AVERAGE |
| 41210853018768_BC | NORMAL | 0.148824779 | AVERAGE | -0.486573454 | AVERAGE |

**Supplementary Table 3:**
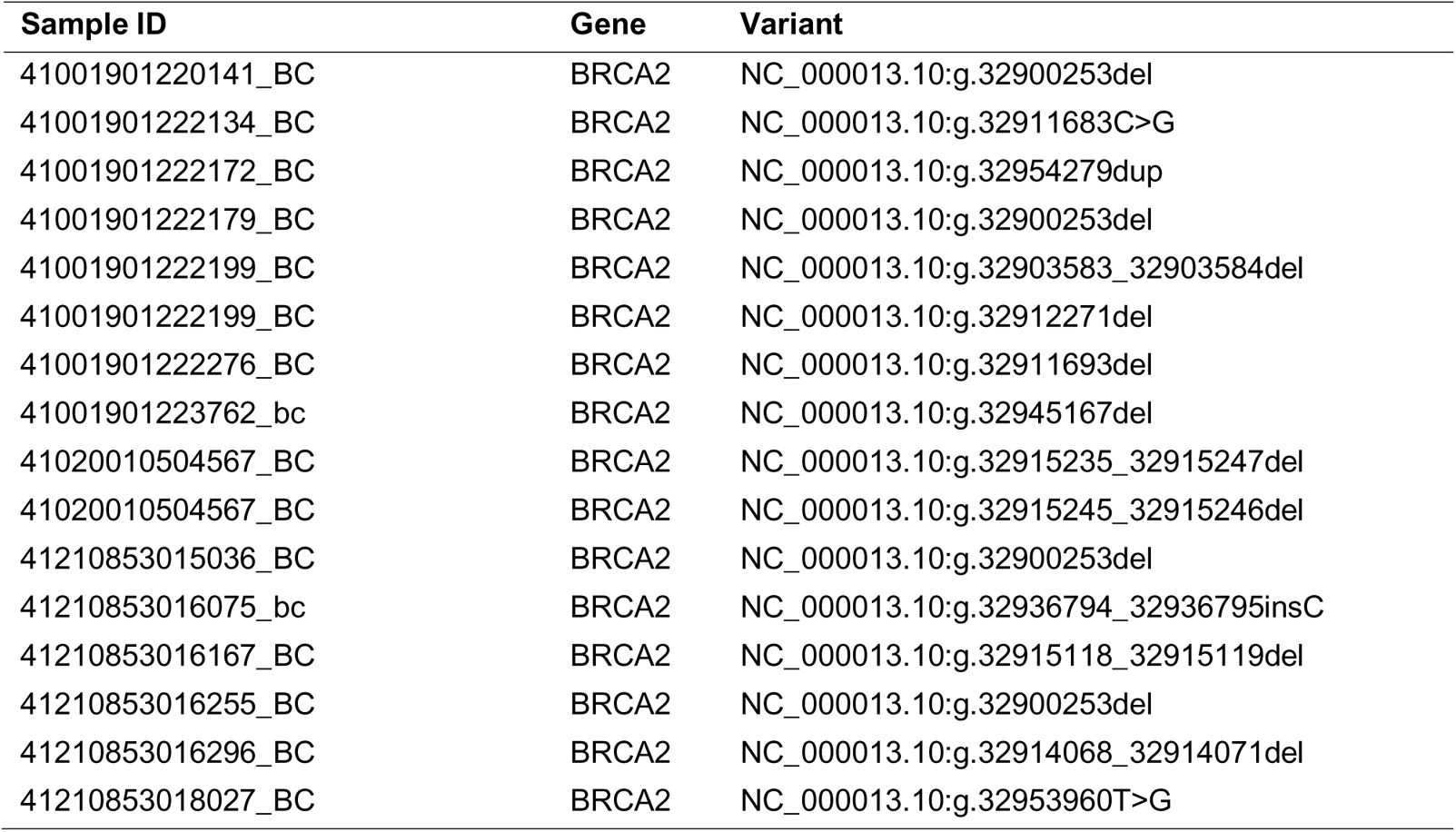
Pathogenic mutations in BRCA1/2 detected in individuals of the study cohort.

**Supplementary Table 4:** Inclusion and exclusion criteria for the clinical validation study.

|  | Control Group | Case Group |
| --- | --- | --- |
| Gender | Female |  |
| Age | 35 – 75 years old |  |
| Breast cancer status | ☐ Had never ever been diagnosed with breast cancer<br>☐ Did not experience any symptoms related to breast cancer<br>☐ Without first-degree relationship with breast cancer case. | ☐ Had ever been diagnosed with breast cancer<br>☐ With or without first-degree relationship with breast cancer case. |
| Ovarian cancer status | Without family history of ovarian cancer | With or without family history of ovarian cancer |

## Notes

### Competing Interest Statement

The authors are affiliated with their respective organizations. BR, SGT, KNR, JA, FA, EAF, MGP, MDV, JH, LHU, YM, KIJ, MA, GG, FS, AF, SSM, LS and AI are affiliated with NalaGenetics. NalaGenetics is a biotech company focusing on personalized medications, diet, and screening. The risk prediction tool investigated in the study was developed by NalaGenetics. FAA, MW, STBS, AW, AS and SJH are affiliated with SJH Initiatives. SJH Initiatives is a research body under MRCCC Siloam Hospitals network whose one of the main focuses is on breast cancer research.
SA is affiliated with Atma Jaya Catholic University of Indonesia. Atma Jaya is a private Catholic university based in Jakarta, Indonesia. The authors have no financial gain or loss in any form that could result from the publication of this manuscript, but the absence of authors without any organizational affiliation could be considered a non-financial competing interest.

### Funding Statement

This study was funded by Nalagenetics.

### Author Declarations

The Medical & Health Research Ethics Commission of MRCCC Hospital gave ethical approval for this work.

### Summary of Updates

A revision has been implemented to enhance the depth of the analysis by integrating discussions concerning pathogenic mutations in BRCA1/2 genes, which serve as exemplary instances of monogenic risk. Additionally, minor textual modifications have been executed, and supplementary references have been included within the document to fortify the research findings.

