## Supplementary File 1 for "A combined risk model shows viability for personalized breast cancer risk assessment in the Indonesian population"

| 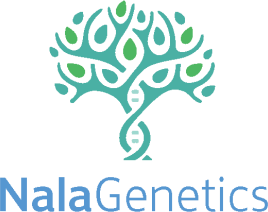 | | | **Specimen Details** | **Lab Barcode Accession No.** | | | |
| --- | --- | --- | --- | --- | --- | --- | --- |
|  |  |  | Specimen Type* : Ordered by* : Consultant :  Clinic’s Name* : Clinic’s Address* : Collection Date* : |  | | | |
| **Patient Details** | | | | | | | |
| **Full Name***  **Date of Birth***  **(DD-MM-YYYY)** |  | : **ID (KTP)***  : **Phone Number*** | | | | :  : (+62) | |
| **Age*** |  | : **Height*** :……………………cm **Weight*** | | | | | :………………………kg |
| **Gender***  **Address*** |  | - Female ☐ Male **Menopausal Status*** : ☐ Pre-menopause ☐ Post-menopause*****   : | | | | | |
|  |  | **Postal Code*** : | | | | | |
| **Ethnicity*** |  | - Ashkenazi Jew ☐ Filipino - Caucasian ☐ Hawaiian - African ☐ Indonesian - Hispanic (US Born) ☐ Other Pacific Islander - Hispanic (Foreign Born) ☐ Other Asian - Chinese ☐ Unknown - Japanese | | | If selecting “Indonesian”, please specify further (e.g., Maluku, Javanese, Sundanese, etc.) | | |
| **General Detail** | | | | | | | |
| Eating / drinking before sample collection: minute / hour (cross out one)  Food / drink before sample collection: | | | | | | | |
| **Assessment Related to Examination (Breast Cancer Risk)** | | | | | | | |
| Age of first menstrual period | | | | - Unknown ☐ 12-13 years old - 7-11 years old ☐ > 13 years old | | | |
| Age at first live birth | | | | - Unknown ☐ 20-24 years old - No births ☐ 25-29 years old - < 20 years old ☐ >= 30 years old | | | |
| Numbers of first degree relative with history of breast cancer (Child, Parent, or Siblings) | | | | - Unknown ☐ 1   ☐ 0 ☐ >1 | | | |
| History of breast biopsy with a benign diagnosis (Fibroadenoma Mammae)  **)****If you have never undergone a biopsy, select ‘Unknown’. If you have undergone biopsy and malignancy or cancer was found, select ‘0’. | | | | - Unknown ☐ 1   ☐ 0 ☐ >1 | | | |
| History of atypical hyperplasia**)****  **)****If you have never undergone a biopsy, select ‘Unknown’. Accumulation of abnormal cells in the breast / Pre-cancer. If you have never had a breast biopsy, please fill in the ‘Unknown’ box. | | | | - Yes ☐ Unknown - No | | | |
| **Relevant Clinical Information (Diagnosis and Medical History)** | | | | | | | |
| Patient has had prior thoracic radiotherapy before age 30 | | | | - Yes ☐ No | | | |
| Patient has had a history of LCIS**)****  **)****Lobular carcinoma in situ (LCIS) is an area (or areas) of abnormal cell growth that increases a person’s risk of developing invasive breast cancer later on in life. | | | | - Yes ☐ No | | | |
| Patient has had a history DCIS**)****  **)****Ductal carcinoma in situ (DCIS) is an area of abnormal cells inside a milk duct in the breast. DCIS is noninvasive, meaning it hasn’t spread out of the milk duct and has a low risk of becoming invasive. | | | | - Yes ☐ No | | | |
| Patient has had a previous diagnosis of breast cancer. | | | | - Yes **)*** ☐ No | | | |

| **)* If Yes:**  Age of the diagnosis:  Estrogen receptor status (If data exists):  Progesterone receptor status (If data exists):  HER2 receptor status (If data exists): | | | | |
| --- | --- | --- | --- | --- |
| Patient has had a second diagnosis of breast cancer | | - Yes | - No |  |
| Patient has had a previous diagnosis of triple-negative breast cancer (ER-negative, PR-negative, HER2-negative) | | - Yes | - No |  |
| **Lifestyle** | | | | |
| How many servings of fruit and vegetables the patient eats a day? | - 1 serving | - 2 – 3 servings | - Once a week or less | |
| How often does the patient exercise per week? | - 150 minutes - 100 minutes | - 60 minutes - 30 minutes | - Unknown | |
| Does the patient smoke? | - Yes | - No |  |  |
| How often does the patient drink alcohol? | - 1-2 drinks a day | - > 3 drinks per day | - Rarely / doesn’t drink alcohol | |
| Use of oral contraceptive | - Never | - Former | - Currently using | |
| Use of hormone replacement therapy | - Never - Former (any type) | - Currently using Estrogen only - Currently using other type | | |
| **Preference** | | | | |
| Preference for risk-reducing therapy | | - Yes | - No |  |
| Preference for lifestyle modification | | - Yes | - No |  |
| Preference for risk-reducing surgery | | - Yes | - No |  |
| Preference for risk-reducing for genetic counseling for multi-gene panel | | - Yes | - No |  |
| **Genetic Predisposition to Breast Cancer (BRCA1/2, p53, PTEN, or other gene mutations)** | | | | |
| Did the patient have any genetic predisposition to breast cancer? | | - Unknown | - Yes | - No |
| How many of the patient’s family relatives have known genetic predisposition to breast cancer? | | - Unknown - (Please fill 0 - 10) - >10 | | |
| **Screening and Monitoring (Part 1)** | | | | |
| When was the last time the patient had breast cancer screening? | - Never - In the last 3 months | - In the last 6 months - 1 year ago | - 2 year ago - > 2 year ago | |
| When was the last time the patient had ovarian cancer screening? | - Never - In the last 3 months | - In the last 6 months - 1 year ago | - 2 years ago - > 2 years ago | |
| **Screening and Monitoring (Part 2)** | | | | |
| Patient had normal results for breast cancer screening according to clinical practice baseline assessment. | - Unknown | - Yes | - No |  |
| Patient had normal results from gynecologic assessment to assess the risk of endometrial cancer. | - Unknown | - Yes | - No |  |
| **Test Ordered** | | | | |
| - Nala Risk Prediction™ | | | | |
| **Deliver To :**  PT Nalagenetik Riset Indonesia  Rukan Permata Senayan Blok H1 H2 Jakarta, Indonesia 12210 | **Notes** | | | |
| *“I have filled out this survey truthfully and to the best of my ability. If the information provided is found to be inaccurate, I understand that it may impact the accuracy and quality of my test results.”* | | | | |
