## Supplementary File 2 for "A combined risk model shows viability for personalized breast cancer risk assessment in the Indonesian population"

### Nala Risk Prediction

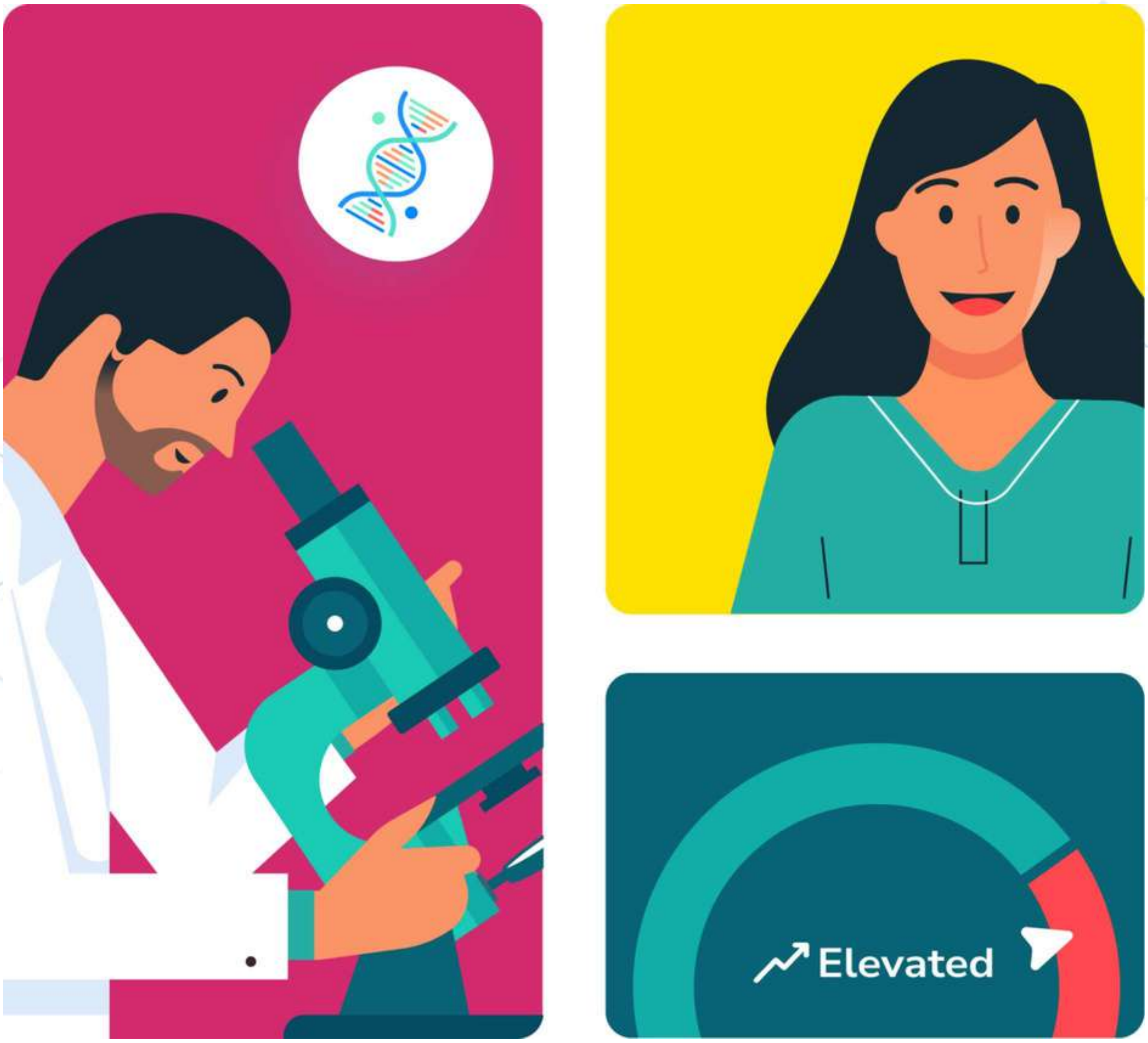


#### ProdTest230224 Patient001

Date of Birth : **01 January 1983**

Report Date : **24 February 2023**

Powered by

NalaGenetics

#### Personal Information

| | |
|---|---|
| Nalagenetics ID | PRPA8324021 |
| Name | ProdTest230224 Patient001 |
| Date of Birth | 01 January 1983 |
| National ID | N/A |
| Gender | Female |

#### Doctor Details

| | |
|---|---|
| Doctor Name | PhysTLPN |
| Doctor ID | 123456789 |
| Contact | +62-81212239000 |
| Order ID | 7259-271640 |

#### Sample Details

| | | | |
|---|---|---|---|
| Clinic Name | TLPN | Lab Address | Jl Pencenogan No.72 |
| Address | Triha Lab | Collected Date | 23 Feb 2023 (06:48 UTC) |
| Type of Sample | Buccal Swab | Received Date | 24 Feb 2023 (07:15 UTC) |
| Test Method | Micro Array | Result Verified | printed in report verification receipt |
| Clinical Director | N/A | | |

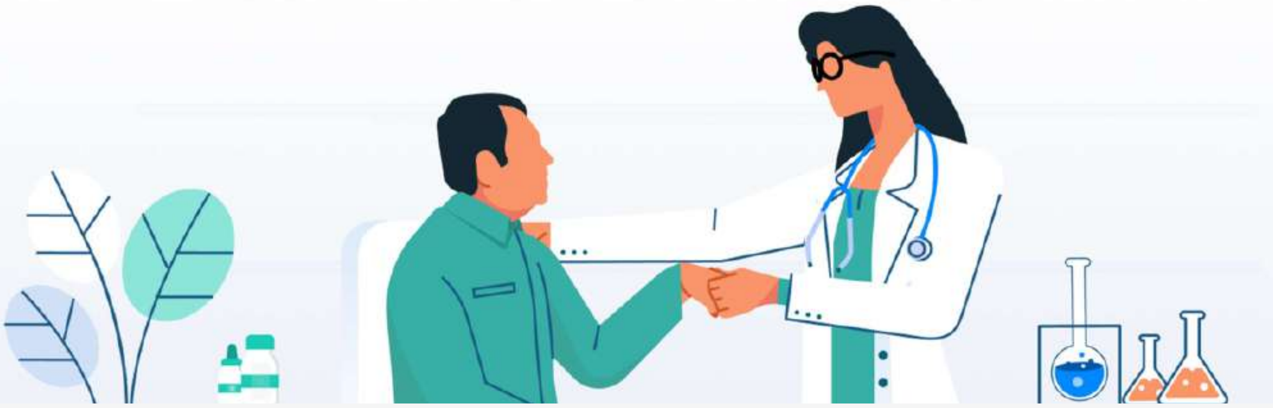

This report was validated and generated automatically. No signature is required. Recommendations given in this report are made using a lab developed test which should not supersede clinical judgement or medical expertise.

### Introduction

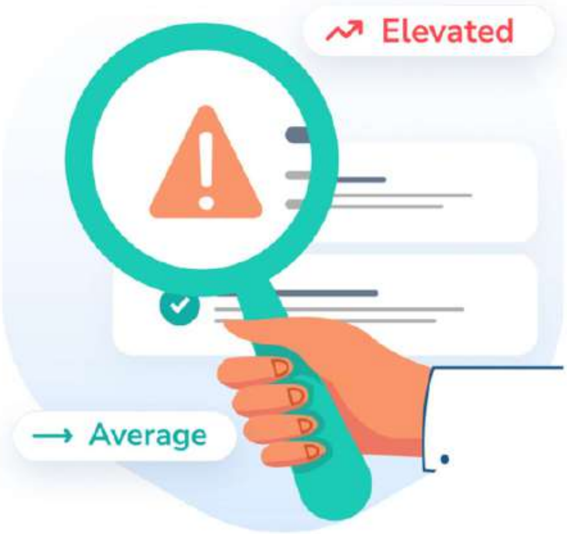


#### What is Nala Risk Prediction report?

Nala Risk Prediction (RP) report is a personalized summary that contains your overall risk towards specific disease and actionable personalized recommendations.

Your Nala RP report is generated as a result of Nala Clinical Decision Support™ analysis which takes into account both your genetic and clinical risk. In addition to your risk profile of being diagnosed to certain disease, Nala RP report also contains your personalized actionnable recommendations

#### Where do we get our recommendations?

Our knowledgebase team collects guidelines and scientific publications with the highest level of evidence to predict a patient's risk of a disease and provide recommendations to prevent the disease. In addition, your personalized actionable recommendations are tailored to your needs based on your overall risk score and categories. Lastly, your report is localized based on your ethnicity and age to provide the most impactful interventions.

The DNA test results are not intended to be used by you as a substitute for professional medical advice. You should always seek the advice of a healthcare provider or physician for any diagnostic purpose with any questions you may have regarding the diagnosis, cure, treatment, mitigation, or prevention of any disease or other medical conditions or impairment or the status of your health.

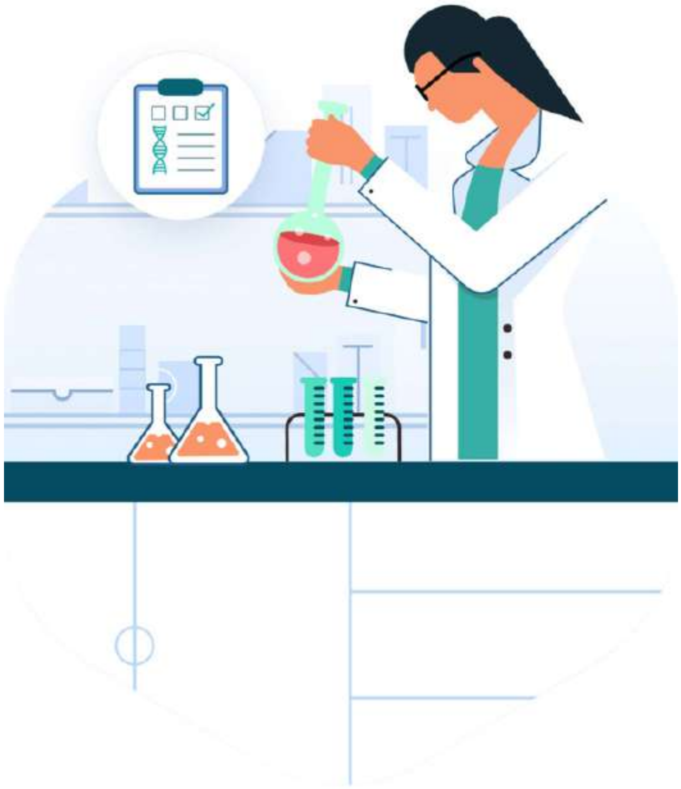

### Disclaimer

The DNA test results are not intended to be used by you as a substitute for professional medical advice. You should always seek the advice of a healthcare provider or physician for any diagnostic purpose with any questions you may have regarding the diagnosis, cure, treatment, mitigation, or prevention of any disease or other medical conditions or impairment or the status of your health.

The laboratory may not be able to process your sample and the laboratory process may result in errors. The laboratory may not be able to process your DNA sample if it does not contain a sufficient amount of DNA. Furthermore, if the DNA sample that you have provided is contaminated and/or corrupted, the accuracy of the DNA test result may be impacted. Even with stringent acceptance criteria for sample processing that meets our high standards, a small, unknown fraction of the data generated during the laboratory process may be un-interpretable or incorrect and may therefore not be able to generate the corresponding report. Should this happen, Nalagenetics will perform imputation to predict the data, instead of using the un-interpretable or incorrect data. You also understand this possibility and will not be entitled to refunds where these occur. For more information on this particular test, you may wish to review the work of Hoetal. 2020 www.ncbi.nlm.nih.gov/pmc/articles/PMC7395776/, which goes into greater detail about the methods/methodology behind the test.

For the avoidance of doubt, Nalagenetics shall only analyze, via genotyping, the parts of your DNA sequence that are relevant to the disease or condition prescribed in the order and consent form you have agreed upon, with such relevance to be determined by Nalagenetics at its sole discretion. You further agree and acknowledge that any relation of the data, predictions, and/or results obtained to other disorders and/or diseases is entirely incidental, and you should seek advice from your healthcare provider regarding the results and their relation to such other disorders and/or diseases (additional charges may apply).

Your sample will be processed with an affiliated laboratory that complies with the prevailing regulations in Indonesia and/or Singapore that implement good clinical laboratory practices. Moreover, any research that Nalagenetics performs complies with IRB-approved protocols, and we treat the data and material that we have received in accordance with the genetic testing as confidential information.

All data and material that have been received by us in accordance with the genetic test are reviewed by a clinical director and/or genetic counselor before being reported by Nalagenetics. There may be differences in the interpretation of the results reported from different laboratories, depending on the study used as references.

Data, results, and/or predictions from Nalagenetics risk assessment are based on information provided by you through your physician at the time of your order, and Nalagenetics shall not be responsible or liable for or on account of any incorrect and or inaccurate information provided by you through the prescribed order form or otherwise, and Nalagenetics shall not be obliged to verify or otherwise look into any information so provided.

You expressly agree and acknowledge that all interpretation, assessment, and/or analysis of the information provided is based on the prevailing knowledge and accepted standards and practices at the time, including but not limited to medical literature, scientific databases, and/or clinical guidelines.

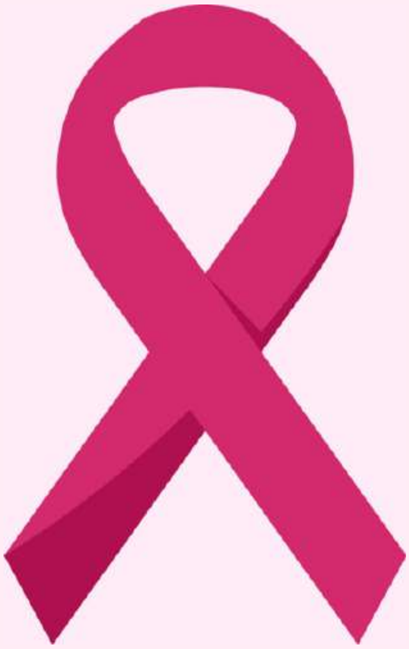

### BREAST CANCER

In 2020, there were 2.3 million women diagnosed with breast cancer and 685 000 deaths globally. As of the end of 2020, there were 7.8 million women alive who were diagnosed with breast cancer in the past 5 years, making it the world's most prevalent cancer.

Breast cancer arises in the lining cells (epithelium) of the ducts (85%) or lobules (15%) in the glandular tissue of the breast. Initially, the cancerous growth is confined to the duct or lobule ("in situ") where it generally causes no symptoms and has minimal potential for spread (metastasis).

Over time, these in situ (stage 0) cancers may progress and invade the surrounding breast tissue (invasive breast cancer) and then spread to the nearby lymph nodes (regional metastasis) or other organs in the body (distant metastasis). If a woman dies from breast cancer, it is because of widespread metastasis.

Breast cancer treatment can be highly effective, especially when the disease is identified early. Treatment of breast cancer often consists of a combination of surgical removal, radiation therapy, and medication (hormonal therapy, chemotherapy, and/or targeted biological therapy) to treat microscopic cancer that has spread from the breast tumor through the blood. Such treatment can prevent cancer growth and spread, thereby saving lives.

*Source:*
*World Health Organization (March 2021)*

### Your Results

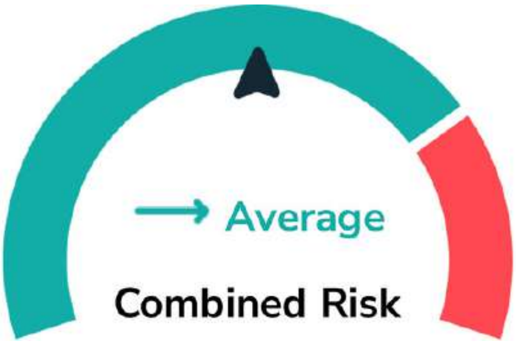


You are at lower chance of developing Breast Cancer in the next 5 years

#### What does this result say about my health?

This is not a diagnosis. Both genetic risk and clinical or lifestyle risk factors contribute to a diagnosis. Genetics only contribute to about 30% of breast cancer risk.

#### How is combined risk defined?

Your combined risk is derived from both your genetic and clinical risk. By comparing your risk scores with thresholds from established guidelines, we determine your risk of being diagnosed with the disease in the next 5 years as “Average” or “Elevated” compared to the population.

| Genetic Risk | Clinical Risk | Combined Risk |
|---|---|---|
| Elevated | Elevated | ↗ Elevated |
| Elevated | Average | ↗ Elevated |
| Average | Elevated | ↗ Elevated |
| Average | Average | → Average |

This report was validated and generated automatically. No signature is required. Recommendations given in this report are made using a lab developed test which should not supersede clinical judgement or medical expertise.

### Your Genetic Risk

#### → Average

5-year absolute risk score

**1.01** %

Current age

**40** y.o

Your risk of breast cancer in the next 5 years based on genetic risk is categorized as Average because your score is lower than 1.7%.

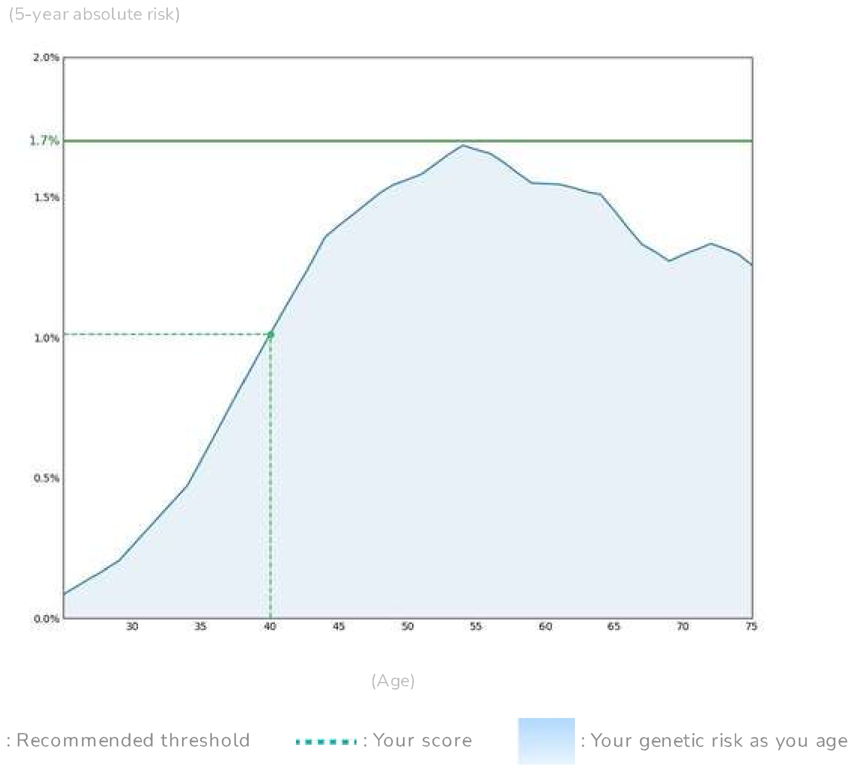


#### How do we get your risk score?

Your 5-year absolute genetic risk score is calculated from your biological sample and genetic analysis, genetic risk score is based on a curated Polygenic Risk Score model for breast cancer. This method takes into account the variant that makes you unique. In addition, the PRS model that we used has been validated in the South East Asian Population.

< **1.7%** Average | ≥ **1.7%** Elevated

*Source:*
*Polygenic Risk Scores for Prediction of Breast Cancer and Breast Cancer Subtypes (PMID: 30554720)*

This report was validated and generated automatically. No signature is required. Recommendations given in this report are made using a lab developed test which should not supersede clinical judgement or medical expertise.

### Your Clinical Risk

→ Average

5-year absolute risk score

0.44 %

Current age

40 y.o.

Your risk of breast cancer in the next 5 year based on clinical risk is categorized as Average because your score is lower than 1.7%. This is based on Surveillance, Epidemiology, and End Results (SEER) registry.

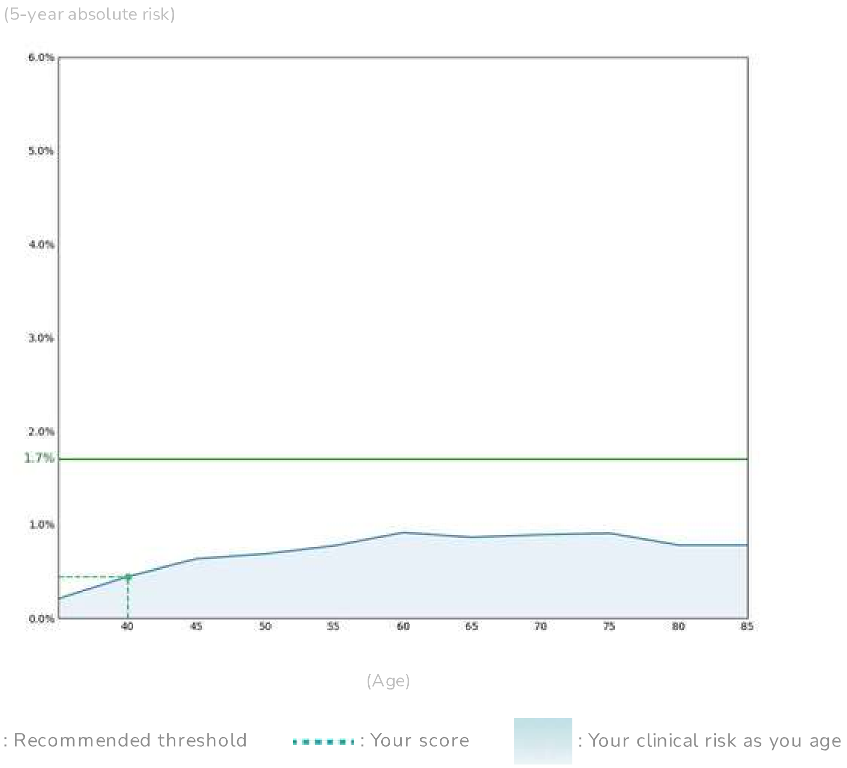


#### How do we get your risk score?

Your 5-year absolute clinical risk score for breast cancer is calculated based on the Gail Model which considers only your medical history and lifestyle information at the point of testing.

< **1.7%** is Average | ≥ **1.7%** is Average

*Source:*
*Polygenic Risk Scores for Prediction of Breast Cancer and Breast Cancer Subtypes (PMID: 30554720)*

This report was validated and generated automatically. No signature is required. Recommendations given in this report are made using a lab developed test which should not supersede clinical judgement or medical expertise.

### What's next?

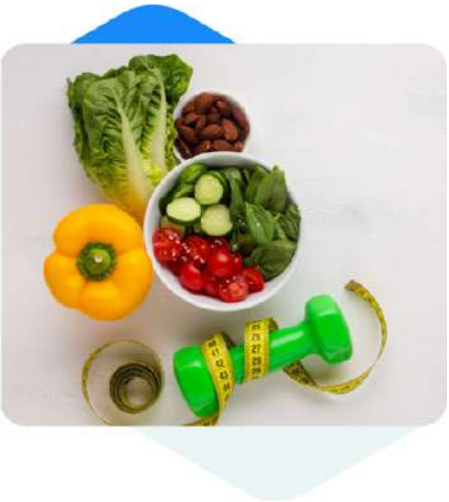

LIFESTYLE

#### Consider healthy habits

Consider building healthy habits to reduce risk.

##### Interesting Facts

- Diet rich in plant-based food, low intake of processed meat and saturated fat has an association with lower risk of breast cancer.
- Lifetime risk of breast cancer is reduced by 9% with increasing amount of physical activity (150-300 minutes of moderate-intensity, physical activity per week).
- Active smoking increases breast cancer risk in women who begins smoking at an early age.

*Source:*


- *Donovan, Micah G. "Do Olive and Fish Oils of the Mediterranean Diet Have a Role in Triple Negative Breast Cancer Prevention and Therapy? An Exploration of Evidence in Cells and Animal Models." Front Nutr. vol. 7, no. 571455, 2020. PMID: 33123546.*
- *Pizot, Cécile. "Physical activity, hormone replacement therapy and breast cancer risk: A meta-analysis of prospective studies." Eur J Cancer, vol. 53, 2016, pp. 138-54, PMID: 26687833.*
- *Gaudet, Mia M. "Active smoking and breast cancer risk: original cohort data and meta-analysis." J Natl Cancer Inst, vol. 105, no. 8, 2013, pp. 515-25, PMID: 23449445.*


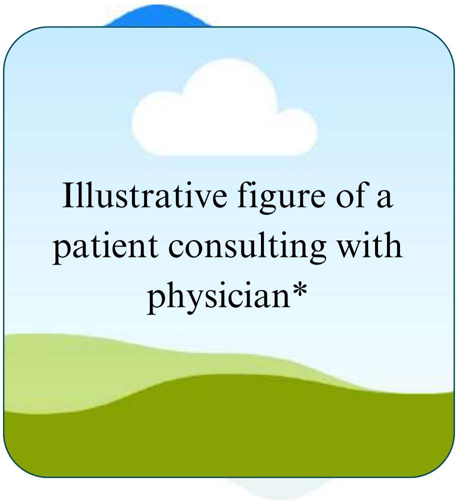


*Please contact the corresponding author to request access to these images or full, unedited versions of the sample reports.

SCREENING AND MONITORING

#### Annual mammography, annual clinical encounter

You may want to speak to your physician for an annual clinical encounter and starting annual mammography screening.

##### Interesting Facts

Mammography screening reduce the estimated breast cancer mortality from up to 9% per year.

*Source:*

*Broeders, Mireille. "The impact of mammographic screening on breast cancer mortality in Europe: a review of observational studies." J Med Screen, vol. 19, no. 1, 2012, pp. 14-25, PMID: 22972807.*

### What's next?

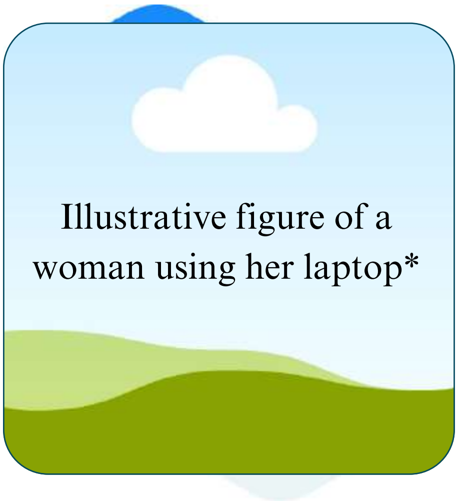


*Please contact the corresponding author to request access to these images or full, unedited versions of the sample reports.

EDUCATION

#### Breast awareness

Learn more about breast cancer self-awareness and management.

#### Interesting Facts

- Carefully performed clinical breast examination (CBE) detects at least 50% of asymptomatic cancer that are potentially curable.
- Personalized videos can increase knowledge of personal risk of breast cancer by 10%.

*Source:*


- *Barton, Mary B. "The rational clinical examination. Does this patient have breast cancer? The screening clinical breast examination: should it be done? How?" JAMA, vol. 282, no. 13, 1999, pp. 1270-80, PMID: 10517431.*
- *Haas, Jennifer S. "Randomized Trial of Personalized Breast Density and Breast Cancer Risk Notification." J Gen Intern Med, vol. 34, no. 4, 2019, pp. 591-597, PMID: 30091121.*

### What's next?

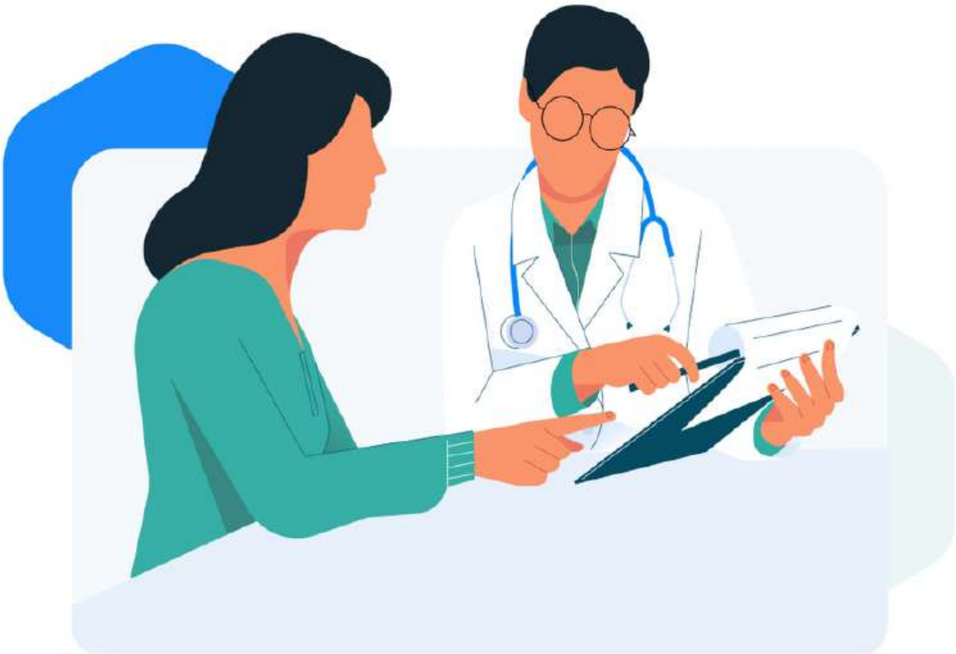

#### Use these recommendations to start a discussion with your doctor

Your survey results have been shared with your doctor. Your doctor will discuss with you the risks and benefits of these options, and the strength of each recommendation based on their level of evidence. These recommendations are curated based on international guidelines, clinical trials, and publications.

#### Do you have questions?

This report was validated and generated automatically. No signature is required. Recommendations given in this report are made using a lab developed test which should not supersede clinical judgement or medical expertise.
