## Supplementary File 3 for "A combined risk model shows viability for personalized breast cancer risk assessment in the Indonesian population"

### Nala Risk Prediction

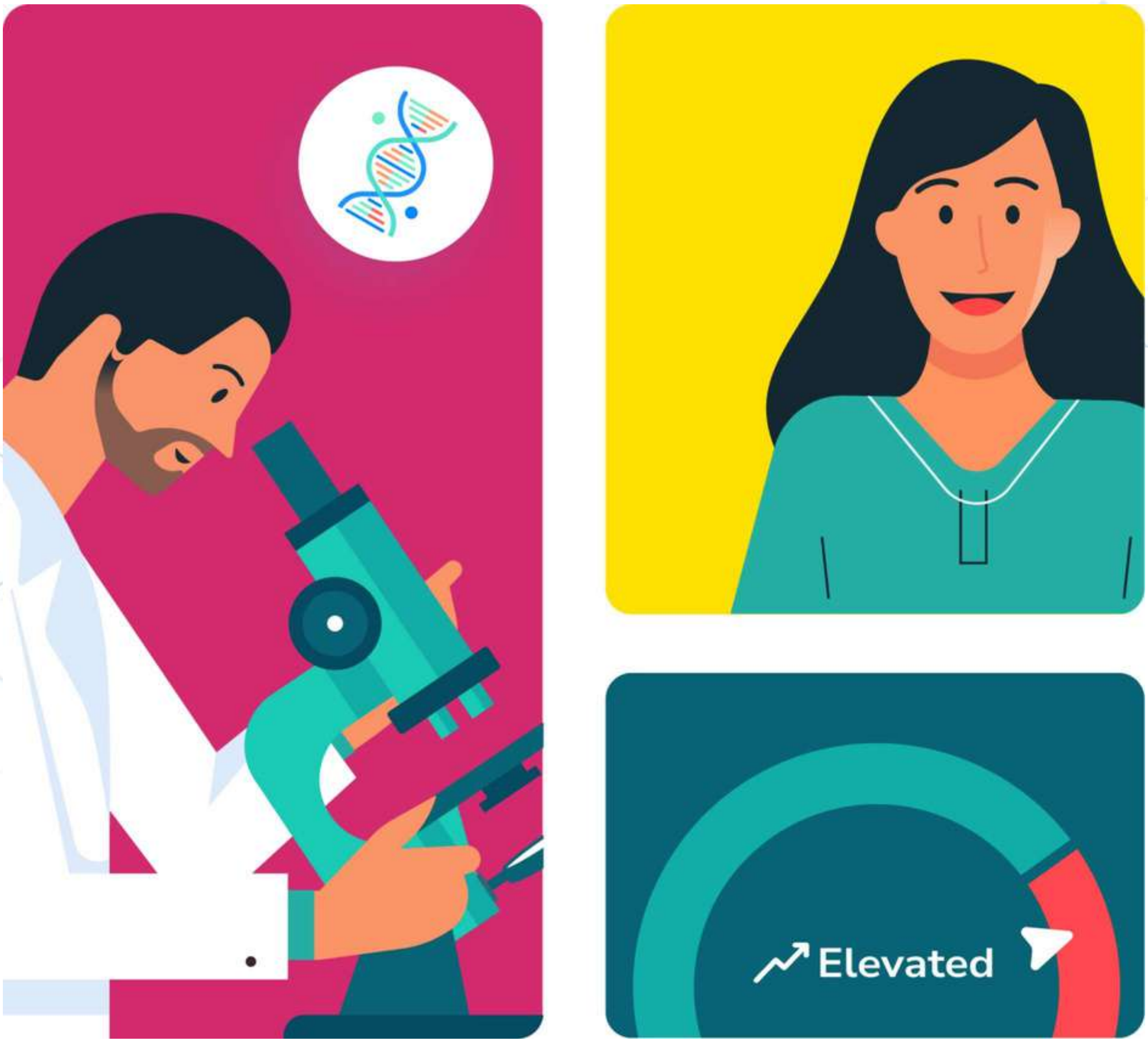


#### UATTest230224 Patient002

Date of Birth : 01 January 1963

Report Date : 24 February 2023

Powered by

NalaGenetics

NalaGenetics

#### Personal Information

| | |
|---|---|
| Nalagenetics ID | **UAPA6324021** |
| Name | **UATTest230224 Patient002** |
| Date of Birth | **01 January 1963** |
| National ID | **N/A** |
| Gender | **Female** |

#### Doctor Details

| | |
|---|---|
| Doctor Name | **PhysTLPN** |
| Doctor ID | **123456789** |
| Contact | **+62-81212239000** |
| Order ID | **3383-259292** |

#### Sample Details

| | |
|---|---|
| Clinic Name | **TLPN** |
| Address | **Triha Lab** |
| Type of Sample | **Buccal Swab** |
| Test Method | **Micro Array** |
| Clinical Director | **N/A** |
| Lab Address | **Jl Pencenogan No.72** |
| Collected Date | **23 Feb 2023 (06:52 UTC)** |
| Received Date | **24 Feb 2023 (07:15 UTC)** |
| Result Verified | **printed in report verification receipt** |

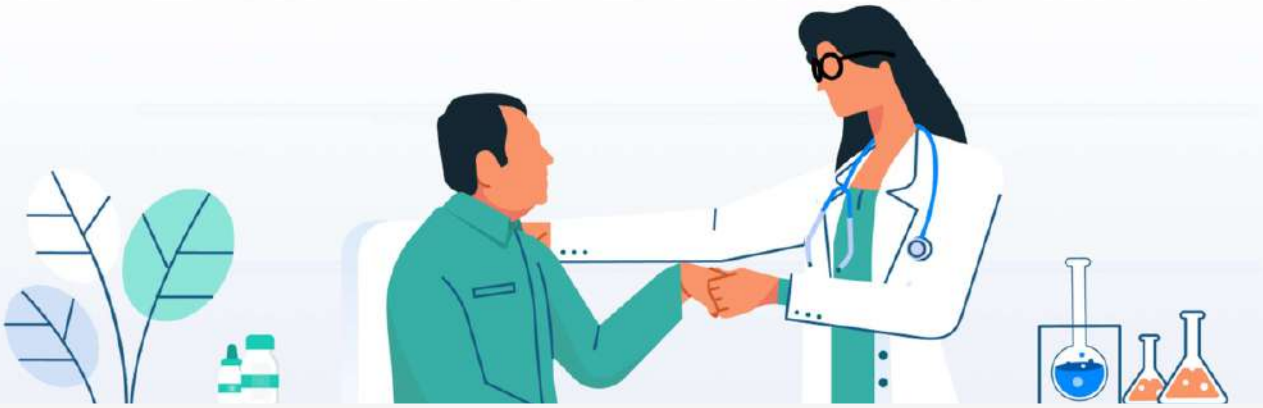

This report was validated and generated automatically. No signature is required. Recommendations given in this report are made using a lab developed test which should not supersede clinical judgement or medical expertise.

### Introduction

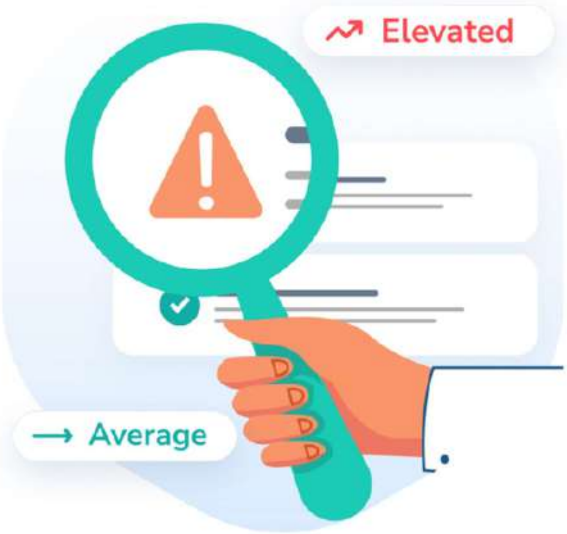


#### What is Nala Risk Prediction report?

Nala Risk Prediction (RP) report is a personalized summary that contains your overall risk towards specific disease and actionnable personalized recommendations.

Your Nala RP report is generated as a result of Nala Clinical Decision Support™ analysis which takes into account both your genetic and clinical risk. In addition to your risk profile of being diagnosed to certain disease, Nala RP report also contains your personalized actionnable recommendations

#### Where do we get our recommendations?

Our knowledgebase team collects guidelines and scientific publications with the highest level of evidence to predict a patient's risk of a disease and provide recommendations to prevent the disease. In addition, your personalized actionable recommendations are tailored to your needs based on your overall risk score and categories. Lastly, your report is localized based on your ethnicity and age to provide the most impactful interventions.

The DNA test results are not intended to be used by you as a substitute for professional medical advice. You should always seek the advice of a healthcare provider or physician for any diagnostic purpose with any questions you may have regarding the diagnosis, cure, treatment, mitigation, or prevention of any disease or other medical conditions or impairment or the status of your health.

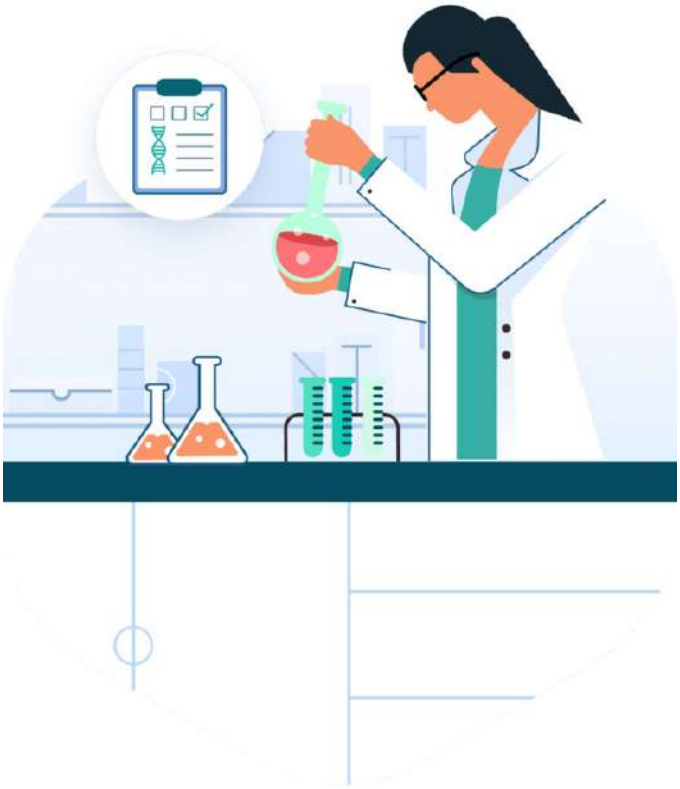

### Disclaimer

The DNA test results are not intended to be used by you as a substitute for professional medical advice. You should always seek the advice of a healthcare provider or physician for any diagnostic purpose with any questions you may have regarding the diagnosis, cure, treatment, mitigation, or prevention of any disease or other medical conditions or impairment or the status of your health.

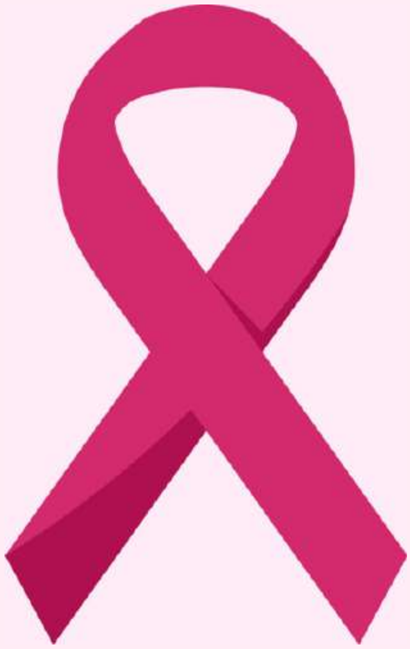

### BREAST CANCER

In 2020, there were 2.3 million women diagnosed with breast cancer and 685 000 deaths globally. As of the end of 2020, there were 7.8 million women alive who were diagnosed with breast cancer in the past 5 years, making it the world's most prevalent cancer.

Breast cancer arises in the lining cells (epithelium) of the ducts (85%) or lobules (15%) in the glandular tissue of the breast. Initially, the cancerous growth is confined to the duct or lobule ("in situ") where it generally causes no symptoms and has minimal potential for spread (metastasis).

Over time, these in situ (stage 0) cancers may progress and invade the surrounding breast tissue (invasive breast cancer) and then spread to the nearby lymph nodes (regional metastasis) or other organs in the body (distant metastasis). If a woman dies from breast cancer, it is because of widespread metastasis.

Breast cancer treatment can be highly effective, especially when the disease is identified early. Treatment of breast cancer often consists of a combination of surgical removal, radiation therapy, and medication (hormonal therapy, chemotherapy, and/or targeted biological therapy) to treat microscopic cancer that has spread from the breast tumor through the blood. Such treatment can prevent cancer growth and spread, thereby saving lives.

*Source:*
*World Health Organization (March 2021)*

### Your Results

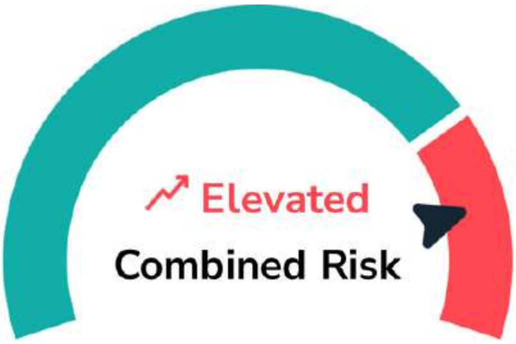


You are at higher chance of developing Breast Cancer in the next 5 years

#### What does this result say about my health?

This is not a diagnosis. Both genetic risk and clinical or lifestyle risk factors contribute to a diagnosis. Genetics only contribute to about 30% of breast cancer risk.

#### How is combined risk defined?

Your combined risk is derived from both your genetic and clinical risk. By comparing your risk scores with thresholds from established guidelines, we determine your risk of being diagnosed with the disease in the next 5 years as “Average” or “Elevated” compared to the population.

| Genetic Risk | Clinical Risk | Combined Risk |
|---|---|---|
| Elevated | Elevated | Elevated |
| Elevated | Average | Elevated |
| Average | Elevated | Elevated |
| Average | Average | Average |

### Your Genetic Risk

**→ Average**

5-year absolute risk score

**0.35** %

Current age

**60** y.o

Your risk of breast cancer in the next 5 years based on genetic risk is categorized as Average because your score is lower than 1.7%.

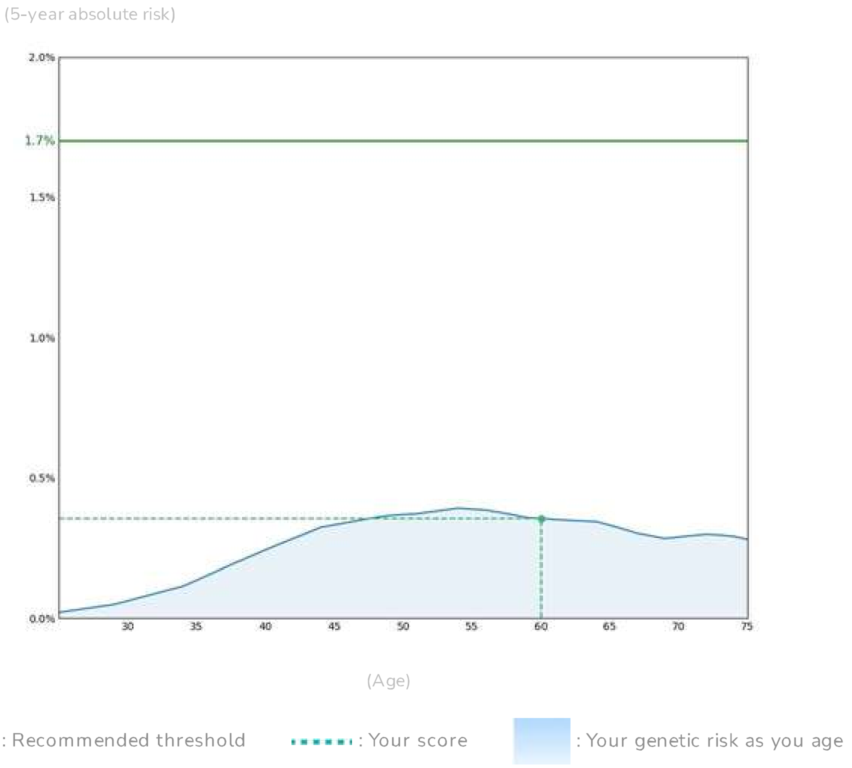


#### How do we get your risk score?

Your 5-year absolute genetic risk score is calculated from your biological sample and genetic analysis, genetic risk score is based on a curated Polygenic Risk Score model for breast cancer. This method takes into account the variant that makes you unique. In addition, the PRS model that we used has been validated in the South East Asian Population.

< **1.7%** Average | ≥ **1.7%** Elevated

*Source:*

*Polygenic Risk Scores for Prediction of Breast Cancer and Breast Cancer Subtypes (PMID: 30554720)*

This report was validated and generated automatically. No signature is required. Recommendations given in this report are made using a lab developed test which should not supersede clinical judgement or medical expertise.

### Your Clinical Risk

**Elevated**

5-year absolute risk score

**4.34** %

Current age

**60** y.o.

Your risk of breast cancer in the next 5 year based on clinical risk is categorized as Elevated because your score is higher than 1.7%. This is based on Surveilance, Epidemiology, and End Results (SEER) registry.

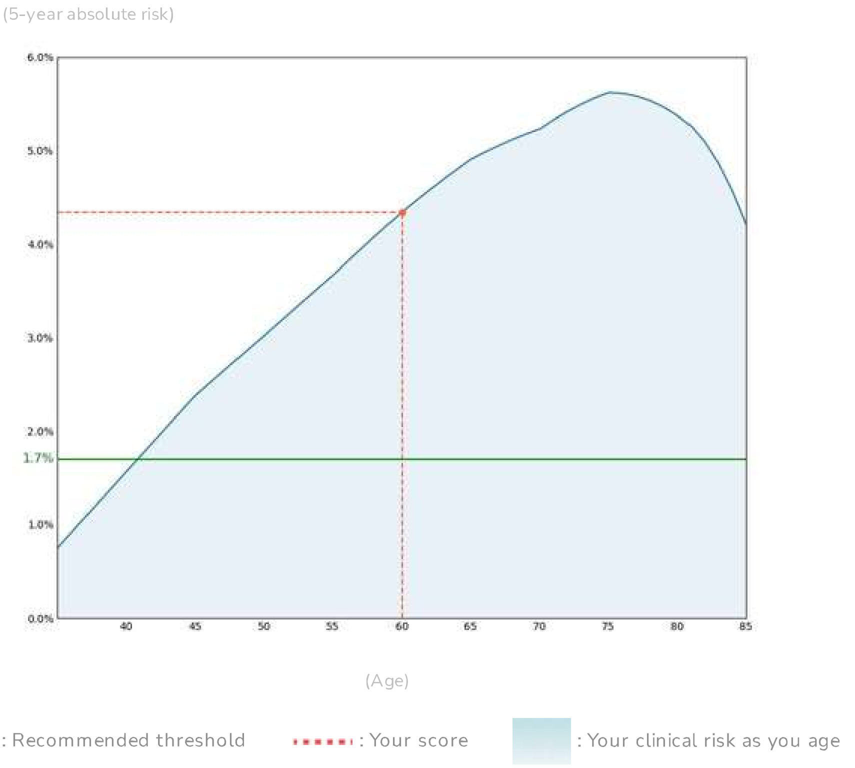


#### How do we get your risk score?

Your 5-year absolute clinical risk score for breast cancer is calculated based on the Gail Model which considers only your medical history and lifestyle information at the point of testing.

< **1.7%** is Average | ≥ **1.7%** is Average

*Source:*
*Polygenic Risk Scores for Prediction of Breast Cancer and Breast Cancer Subtypes (PMID: 30554720)*

This report was validated and generated automatically. No signature is required. Recommendations given in this report are made using a lab developed test which should not supersede clinical judgement or medical expertise.

### What's next?

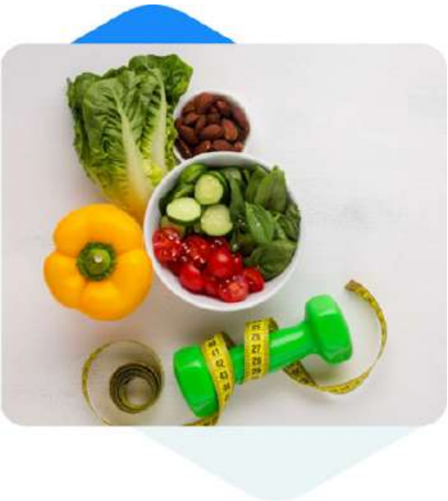

#### LIFESTYLE

##### Consider healthy habits

Consider building healthy habits to reduce risk.

##### Interesting Facts

- Diet rich in plant-based food, low intake of processed meat and saturated fat has an association with lower risk of breast cancer.
- Lifetime risk of breast cancer is reduced by 9% with increasing amount of physical activity (150-300 minutes of moderate-intensity, physical activity per week).
- Active smoking increases breast cancer risk in women who begins smoking at an early age.

*Source:*

- *Donovan, Micah G. "Do Olive and Fish Oils of the Mediterranean Diet Have a Role in Triple Negative Breast Cancer Prevention and Therapy? An Exploration of Evidence in Cells and Animal Models." Front Nutr. vol. 7, no. 571455, 2020. PMID: 33123546.*
- *Pizot, Cécile. "Physical activity, hormone replacement therapy and breast cancer risk: A meta-analysis of prospective studies." Eur J Cancer, vol. 53, 2016, pp. 138-54, PMID: 26687833.*
- *Gaudet, Mia M. "Active smoking and breast cancer risk: original cohort data and meta-analysis." J Natl Cancer Inst, vol. 105, no. 8, 2013, pp. 515-25, PMID: 23449445.*

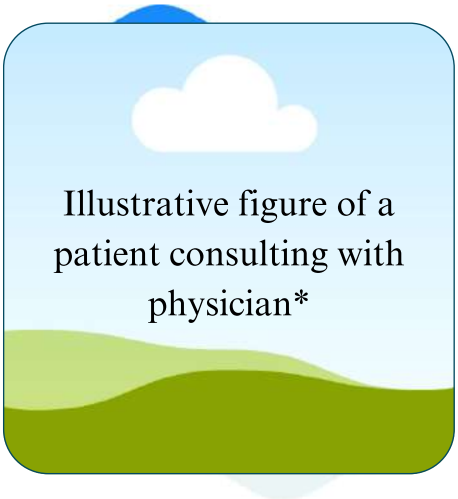


*Please contact the corresponding author to request access to these images or full, unedited versions of the sample reports.

#### SCREENING AND MONITORING

##### Clinical encounter every 6-12 months, annual mammography

You may want to speak to your physician for a clinical encounter every 6-12 months and starting annual mammography screening.

##### Interesting Facts

Mammography screening reduce the estimated breast cancer mortality from up to 9% per year.

*Source:*
*Broeders, Mireille. "The impact of mammographic screening on breast cancer mortality in Europe: a review of observational studies." J Med Screen, vol. 19, no. 1, 2012, pp. 14-25, PMID: 22972807.*

### What's next?

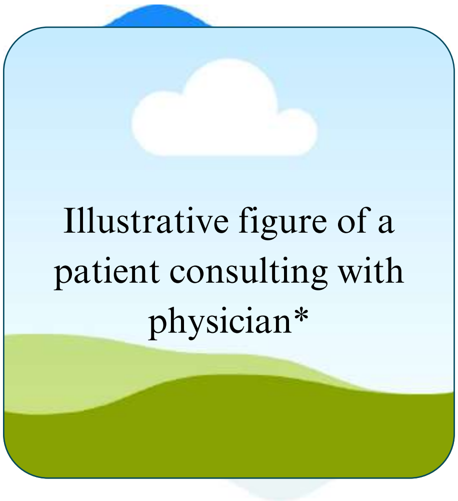


*Please contact the corresponding author to request access to these images or full, unedited versions of the sample reports.

SURGERY

#### Genetic counseling

Consider speaking to a genetic counselor for surgical options.

##### Interesting Facts

- Studies show 50% reduction in breast cancer risk associated with risk reducing salpingo oophorectomy in women who carry mutations in BRCA1/2.
- Studies provide support for concluding that risk-reducing mastectomy provides a high degree of protection against breast cancer in women with a BRCA1/2 mutation.

*Source:*


- *Rebbeck, Timothy R. "Prophylactic oophorectomy in carriers of BRCA1 or BRCA2 mutations." N Engl J Med, vol. 346, no. 21, 2002, pp. 1616-22, PMID: 12023993.*
- *Rebbeck, Timothy R. "Meta-analysis of risk reduction estimates associated with risk-reducing salpingo-oophorectomy in BRCA1 or BRCA2 mutation carriers." J Natl Cancer Inst, vol. 101, no. 2, 2009, pp.80-7, PMID: 19141781.*


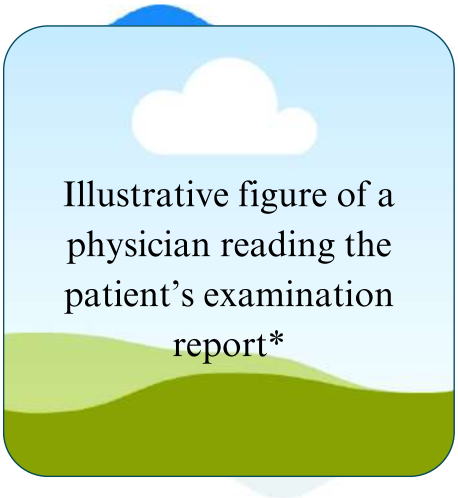


*Please contact the corresponding author to request access to these images or full, unedited versions of the sample reports.

THERAPY

#### Selective estrogen receptor modulator

Speak to your physician about risks and benefits of taking risk-reducing therapy.

##### Interesting Facts

- Selective estrogen receptor modulators (SERMs) reduces overall breast cancer incidence by 38%.
- However, thromboembolic events were significantly increased with all SERMs.

*Source:*


- *Cuzick, Jack. "Selective oestrogen receptor modulators in prevention of breast cancer: an updated meta-analysis of individual participant data." Lancet, vol. 381, no. 9880, 2013, pp. 1827-34, PMID: 23639488.*
- *Cuzick, J. "First results from the International Breast Cancer Intervention Study (IBIS-I): a randomised prevention trial." Lancet, vol. 360, no. 9336, 2002, pp. 817-24, PMID: 12243915.*


This report was validated and generated automatically. No signature is required. Recommendations given in this report are made using a lab developed test which should not supersede clinical judgement or medical expertise.

### What's next?

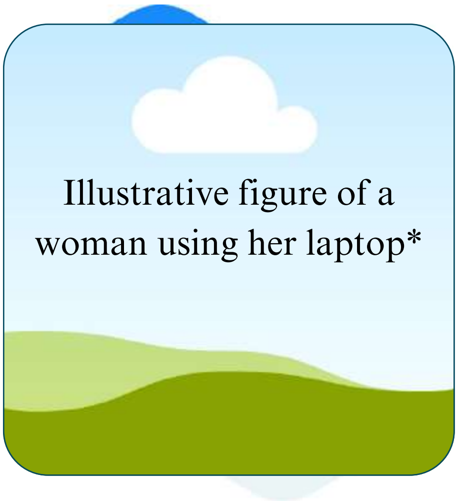


*Please contact the corresponding author to request access to these images or full, unedited versions of the sample reports.

EDUCATION

#### Breast awareness

Learn more about breast cancer self-awareness and management.

##### Interesting Facts

- Carefully performed clinical breast examination (CBE) detects at least 50% of asymptomatic cancer that are potentially curable.
- Personalized videos can increase knowledge of personal risk of breast cancer by 10%.

*Source:*

- *Barton, Mary B. "The rational clinical examination. Does this patient have breast cancer? The screening clinical breast examination: should it be done? How?" JAMA, vol. 282, no. 13, 1999, pp. 1270-80, PMID: 10517431.*
- *Haas, Jennifer S. "Randomized Trial of Personalized Breast Density and Breast Cancer Risk Notification." J Gen Intern Med, vol. 34, no. 4, 2019, pp. 591-597, PMID: 30091121.*

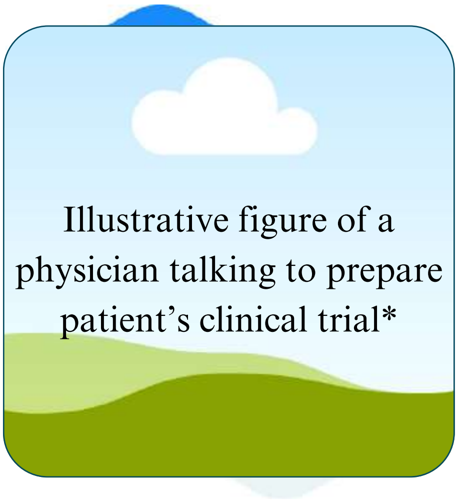


*Please contact the corresponding author to request access to these images or full, unedited versions of the sample reports.

CLINICAL TRIAL

#### Clinical trial

There may be clinical trials available for you to join.

##### Interesting Facts

### What's next?

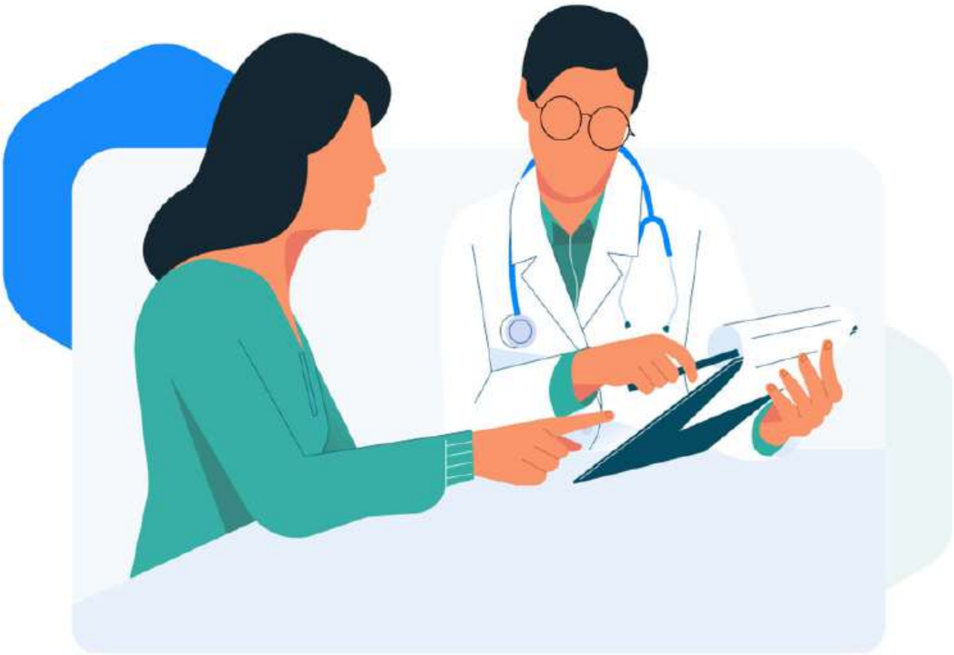

#### Use these recommendations to start a discussion with your doctor

Your survey results have been shared with your doctor. Your doctor will discuss with you the risks and benefits of these options, and the strength of each recommendation based on their level of evidence. These recommendations are curated based on international guidelines, clinical trials, and publications.

#### Do you have questions?

This report was validated and generated automatically. No signature is required. Recommendations given in this report are made using a lab developed test which should not supersede clinical judgement or medical expertise.
